# Stratifying Lung Adenocarcinoma Risk with Multi-ancestry Polygenic Risk Scores in East Asian Never-Smokers

**DOI:** 10.1101/2024.06.26.24309127

**Authors:** Batel Blechter, Xiaoyu Wang, Juncheng Dai, Christiana Karsonaki, Jianxin Shi, Kouya Shiraishi, Jiyeon Choi, Keitaro Matsuo, Tzu-Yu Chen, Rayjean J Hung, Kexin Chen, Xiao-Ou Shu, Young Tae Kim, Parichoy Pal Choudhury, Jacob Williams, Maria Teresa Landi, Dongxin Lin, Wei Zheng, Zhihua Yin, Baosen Zhou, Jiucun Wang, Wei Jie Seow, Lei Song, I-Shou Chang, Wei Hu, Li-Hsin Chien, Qiuyin Cai, Yun-Chul Hong, Hee Nam Kim, Yi-Long Wu, Maria Pik Wong, Brian Douglas Richardson, Shilan Li, Tongwu Zhang, Charles Breeze, Zhaoming Wang, Bryan A Bassig, Jin Hee Kim, Demetrius Albanes, Jason YY Wong, Min-Ho Shin, Lap Ping Chung, Yang Yang, Hong Zheng, Hongji Dai, Yasushi Yatabe, Xu-Chao Zhang, Young-Chul Kim, Neil E Caporaso, Jiang Chang, James Chung Man Ho, Yataro Daigo, Yukihide Momozawa, Yoichiro Kamatani, Masashi Kobayashi, Kenichi Okubo, Takayuki Honda, H Dean Hosgood, Hideo Kunitoh, Shun-ichi Watanabe, Yohei Miyagi, Shingo Matsumoto, Hidehito Horinouchi, Masahiro Tsuboi, Ryuji Hamamoto, Koichi Goto, Atsushi Takahashi, Akiteru Goto, Yoshihiro Minamiya, Megumi Hara, Yuichiro Nishida, Kenji Takeuchi, Kenji Wakai, Koichi Matsuda, Yoshinori Murakami, Kimihiro Shimizu, Hiroyuki Suzuki, Motonobu Saito, Yoichi Ohtaki, Kazumi Tanaka, Tangchun Wu, Fusheng Wei, Mitchell J Machiela, Yeul Hong Kim, In-Jae Oh, Victor Ho Fun Lee, Gee-Chen Chang, Kuan-Yu Chen, Wu-Chou Su, Yuh-Min Chen, Adeline Seow, Jae Yong Park, Sun-Seog Kweon, Yu-Tang Gao, Jianjun Liu, Ann G Schwartz, Richard Houlston, Ivan P Gorlov, Xifeng Wu, Ping Yang, Stephen Lam, Adonina Tardon, Chu Chen, Stig E Bojesen, Mattias Johansson, Angela Risch, Heike Bickeböller, Bu-Tian Ji, H-Erich Wichmann, David C Christiani, Gad Rennert, Susanne M Arnold, Paul Brennan, James McKay, John K Field, Michael P.A. Davies, Sanjay S Shete, Loïc Le Marchand, Geoffrey Liu, Angeline S Andrew, Lambertus A Kiemeney, Shan Zienolddiny-Narui, Kjell Grankvist, Angela Cox, Fiona Taylor, Jian-Min Yuan, Philip Lazarus, Matthew B Schabath, Melinda C Aldrich, Hyo-Sung Jeon, Shih Sheng Jiang, Chung-Hsing Chen, Chin-Fu Hsiao, Zhibin Hu, Laura Burdett, Meredith Yeager, Amy Hutchinson, Belynda Hicks, Jia Liu, Sonja I Berndt, Wei Wu, Junwen Wang, Yuqing Li, Jin Eun Choi, Kyong Hwa Park, Sook Whan Sung, Chang Hyun Kang, Wen-Chang Wang, Jun Xu, Peng Guan, Wen Tan, Chong-Jen Yu, Gong Yang, Alan Dart Loon Sihoe, Yi Young Choi, In Kyu Park, Hsiao-Han Hung, Roel C.H. Vermeulen, Iona Cheng, Junjie Wu, Fang-Yu Tsai, John K.C. Chan, Jihua Li, Hsien-Chih Lin, Jie Liu, Bao Song, Norie Sawada, Taiki Yamaji, Kathleen Wyatt, Hongxia Ma, Meng Zhu, Yifan Wang, Tianchen Qi, Xuelian Li, Yangwu Ren, Ann Chao, Motoki Iwasaki, Junjie Zhu, Guoping Wu, Chih-Yi Chen, Chien-Jen Chen, Pan-Chyr Yang, Victoria L. Stevens, Joseph F. Fraumeni, Kuang Lin, Robin G Walters, Zhengming Chen, Nilanjan Chatterjee, Olga Y Gorlova, Christopher I Amos, Hongbing Shen, Chao Agnes Hsiung, Stephen J Chanock, Nathaniel Rothman, Takashi Kohno, Qing Lan, Haoyu Zhang

## Abstract

**Background:** Lung adenocarcinoma (LUAD) in never-smokers is a major public health burden, especially among East Asian women. Polygenic risk scores (PRSs) are promising for risk stratification but are primarily developed in European-ancestry populations. We aimed to develop and validate single- and multi-ancestry PRSs for East Asian never-smokers to improve LUAD risk prediction.

**Methods:** PRSs were developed using genome-wide association study summary statistics from East Asian (8,002 cases; 20,782 controls) and European (2,058 cases; 5,575 controls) populations. Single-ancestry models included PRS-25, PRS-CT, and LDpred2; multi-ancestry models included LDpred2+PRS-EUR128, PRS-CSx, and CT-SLEB. Performance was evaluated in independent East Asian data from the Female Lung Cancer Consortium (FLCCA) and externally validated in the Nanjing Lung Cancer Cohort (NJLCC). We assessed predictive accuracy via AUC, with 10-year and (age 30-80) absolute risks estimates.

**Results:** The best multi-ancestry PRS, using East Asian and European data via CT-SLEB (clumping and thresholding, super learning, empirical Bayes), outperformed the best East Asian-only PRS (LDpred2; AUC=0.629, 95% CI:0.618,0.641), achieving an AUC of 0.640 (95% CI:0.629,0.653) and odds ratio of 1.71 (95% CI:1.61,1.82) per SD increase. NJLCC Validation confirmed robust performance (AUC =0.649, 95% CI: 0.623, 0.676). The top 20% PRS group had a 3.92-fold higher LUAD risk than the bottom 20%. Further, the top 5% PRS group reached a 6.69% lifetime absolute risk. Notably, this group reached the average population 10-year LUAD risk at age 50 (0.42%) by age 41, nine years earlier.

**Conclusions:** Multi-ancestry PRS approaches enhance LUAD risk stratification in East Asian never-smokers, with consistent external validation, suggesting future clinical utility.

## Introduction

Lung cancer is a major global health challenge, responsible for about 20% of all cancer deaths in 2020^1^. While smoking is the primary etiologic factor, around 25% of lung cancer cases occur in never-smokers, with significant geographical variations^2^. Notably, in East Asia, never-smoking women exhibit high incidences of lung adenocarcinoma (LUAD), the predominant histologic subtype^3^.

Beyond smoking, risk factors for LUAD include environmental and occupational exposures, lifestyle, family history, and genetic susceptibility^4–7^. Genome-wide association studies (GWAS) for never-smoking lung cancer across East Asian and European (EUR) populations have identified 28 susceptibility loci at 25 independent regions^8–13^. These GWAS findings enhance our understanding of LUAD’s genetic architecture; yet, the translation of these findings into clinical applications requires further investigation.

Polygenic risk scores (PRS) aggregate the effects of individual common single nucleotide polymorphisms (SNPs) to estimate genetic risk for diseases, potentially playing a role in cancer prevention and screening. Previous efforts primarily utilized European populations for lung cancer PRS development^14,15^ to better identify high-risk individuals. Nonetheless, because of differences in linkage disequilibrium and allele frequencies across ancestries, such PRSs often underperform in East Asian never-smokers, reflecting the need to address the disproportionate burden of LUAD in this population and to advance the integration of PRS into risk stratification for lung cancer prevention and screening. Despite ongoing efforts to integrate PRS into clinical practices for complex diseases, as seen in initiatives like the electronic Medical Records and Genomics (eMERGE) network^16^, Veterans Affairs Genomic Medicine at Veteran Affairs (GenoVA) study^17^, and the Women Informed to Screen Depending On Measures of risk (WISDOM) study^18^, lung cancer has been excluded. The predominant global burden of lung cancer, driven primarily by smoking, overlooks the significant impact and potential benefits of PRS in never-smoking populations.

The recent expansion of GWAS across diverse populations, combined with advancements in PRS methodology, underscores a pivotal shift towards enhancing disease outcome prediction beyond European populations^19^. Notably, the development of multi-ancestry PRS methods marks a considerable advancement, leveraging data from various ancestral backgrounds to enrich the predictive accuracy and robustness of PRSs^20–26^. By jointly modeling genetic data from multiple populations, these approaches improve the predictive power of PRSs, thereby enhancing disease outcome predictions in non-European populations.

While the development and validation of PRSs are critical, the application of an established PRS in estimating the absolute risk of a disease offers valuable potential for risk stratification, potentially guiding clinical interventions, such as lung cancer screening. Moreover, projecting necessary sample sizes for future studies to achieve varying levels of PRS predictive accuracy is an underexplored area that could significantly impact epidemiological research design and cost-effectiveness.

This study aims to evaluate the predictive performance of single- and multi-ancestry PRSs for LUAD in never-smoking East Asian individuals using state-of-the-art methodologies. By integrating multiple datasets, we constructed PRSs based on summary statistics for 5,622 never-smoking cases and 21,813 never-smoking controls from East Asian and European ancestries. We assessed the performance of these PRSs using independent, individual level data of 4,438 never-smoking East Asian cases and 4,544 never-smoking East Asian controls.

Furthermore, we estimated the lifetime and 10-year absolute risks of LUAD using the most accurate PRSs developed. Lastly, we projected the sample sizes needed in future research to achieve specific levels of prediction accuracy with PRSs in East Asian never-smokers (**Figure 1**).

**Figure 1.**
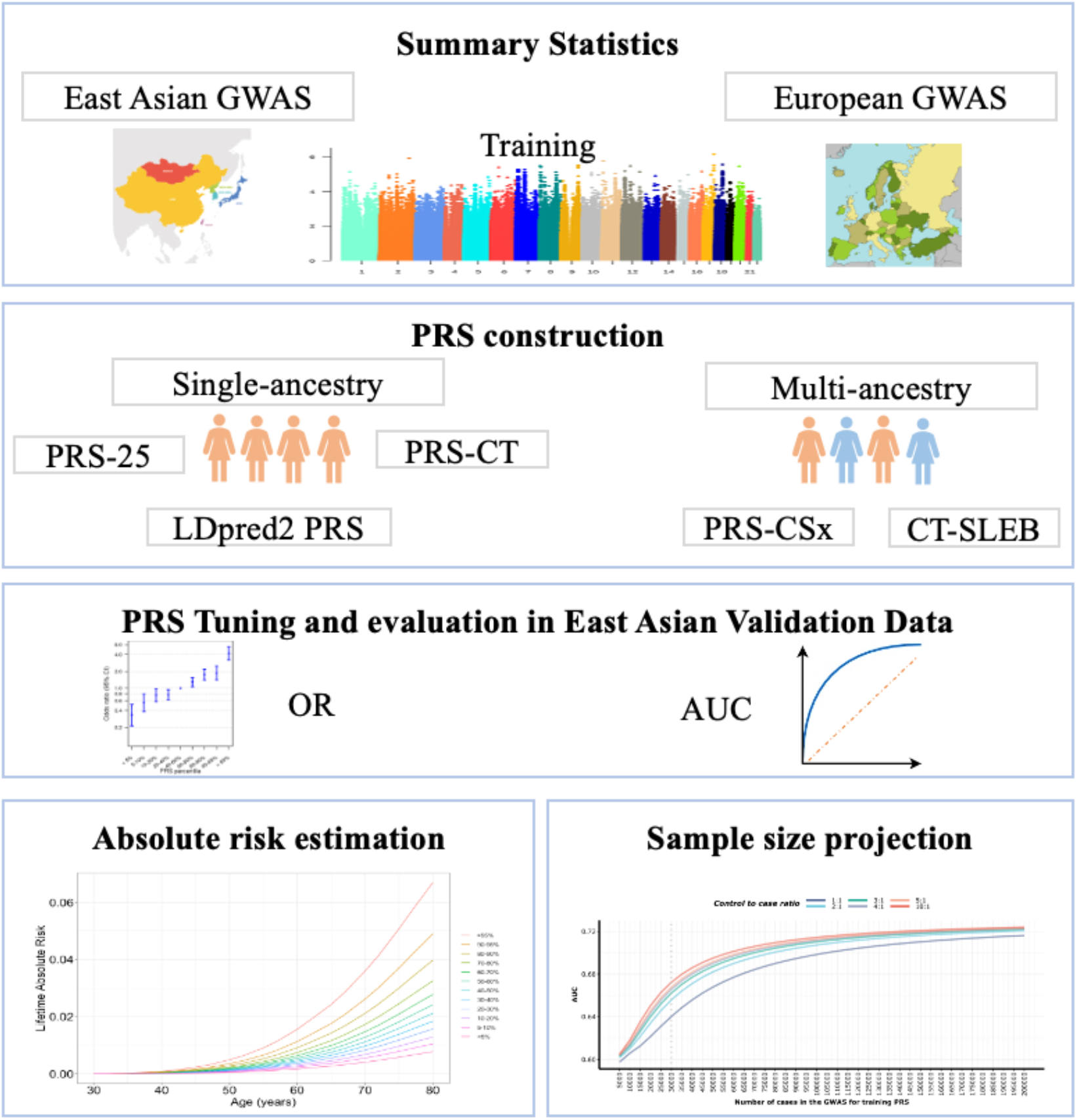
Overview of data structure, polygenic risk score (PRS) development, validation and application. Summary statistics from East Asian genome-wide association studies (GWAS) were used to develop single-ancestry PRS using methods such as a simple PRS constructed using 25 SNPs that have previously reached genome-wide significance (i.e., P<5×10^−8^) (PRS-25), a PRS using the clumping and thresholding (CT) method (PRS-CT) incorporating 8 SNPs, and a PRS using a genome-wide Bayesian-based approach, LDpred2 (LDpred2 PRS) incorporating close to a million SNPs. For the multi-ancestry PRS development, we also used summary statistics from European (EUR) GWAS, applying the PRS-CSx method that leveraged genome-wide association summary statistics for close to a million SNPs with a Bayesian continuous shrinkage prior to model SNP effect sizes across populations, as well as CT-SLEB method, which enhances the standard CT methods with a two-dimensional approach to select SNPS for East Asian PRS construction by incorporating over 2 million SNPs. Tuning and validation of each PRS was conducted in an independent East Asian individual-level data. Relative risk per PRS quantile was calculated as an odds ratio (OR) with the middle quantile (40^th^ to 60^th^ percentile) set as the reference, and the area under the receiver operating curve (AUC) was estimated for each PRS. CT-SLEB PRS was used to estimate 10-year and lifetime cumulative absolute risk, and PRS-CT and PRS-LDpred2 were used for sample size projection.

## Methods

### Study design and data sources

#### Training data compilation: study population and genotyping

The studies, genotyping protocols, and quality control for PRS construction have been previously described in detail^27^. Briefly, biological sex and main continental ancestry were genetically determined for all participants in each study. East Asian single-ancestry PRSs for LUAD were constructed using summary data from 3,564 never-smoking LUAD cases and 16,238 never-smoking controls of East Asian ancestry from the Nanjing Lung Cancer Cohort (NJLCC)^28–31^, National Cancer Center of JAPAN (NCC), and the Research Institute and Aichi Cancer Center (ACC). The NJLCC study combined data from several cities, genotyped by Affymetrix Genome-Wide Human SNP Array 6.0, Illumina OncoArray, Illumina Asian Screening Array, as well as whole genome-sequecing by Illumina HiSeq or NovaSeq platforms^28–31^.The NCC study included lung cancer patients and control data from multiple Japanese studies, with genotyping conducted on Illumina HumanOmniExpress and HumanOmni1-Quad genotyping platforms. Similarly, the ACC study pooled data from multiple Japanese medical institutions and the Nagahama Study, with genotyping performed using Illumina 610k and Illumina660k platforms. Imputation and quality control were conducted using the pipeline described in Shi et al. (2023)^27^. SNPs with minor allele frequency < 0.01, Hardy-Weinberg Equilibrium p-value 1.0 × 10^−6^, or imputation quality scores < 0.5 were excluded. Genotype imputation was performed using IMPUTE2 and the 1000 Genomes Project East Asian samples (Phase 3) as the reference panel. Variants present in the training datasets (NJLCC, NCC, and ACC) but not in the validation dataset (FLCCA) were excluded from analyses. To construct the multi-ancestry PRS, we further incorporated GWAS summary statistics from 2,058 never-smoking LUAD cases and 5,575 never-smoking controls of European ancestry, genotyped using Illumina Infinium OmniExpress-24 v1.2 BeadChips and Illumina Human660W-Quad BeadChip^13^. Weights for all East Asian single-ancestry PRSs and the East Asian portion of the multi-ancestry PRSs were estimated from a meta-analysis restricted to never-smokers from NJLCC, NCC and ACC studies.

#### Tuning and validation: study population and genotyping

For PRS tuning and internal validation, we used 4,438 never-smoking LUAD cases and 4,544 never-smoking controls from the Female Lung Cancer Consortium in Asia (FLCCA)^10,11^, an international consortium composed of never-smoking East Asian women from regions including Mainland China, Singapore, Taiwan, Hong Kong, South Korea and Japan ^10,11^. All samples were genotyped using the Illumina 660W, 370K or 610Q microarrays. Biological sex and ancestry were genetically determined, and information on age and family history of lung cancer were collected through a questionnaire. The FLCCA data were randomly and equally divided for tuning (2,219 cases, 2,272 controls) and validation (2,219 cases, 2,272 controls).

We externally validated the PRSs in two independent studies, including the NJLCC-WGS^29^ and the China Kadoorie Biobank (CKB). The NJLCC-WGS is a subproject of NJLCC, aiming to identify rare variants of lung cancer by whole-genome sequencing. In the NJLCC-WGS, 1,643 never-smoking LUAD cases (473 male; 1,170 female) and 2,235 never-smoking controls (795 male; 1,440 female) were sequenced by the Illumina HiSeq or NovaSeq platforms. The alignment, genotyping and quality control are previously described in detail^29^. Briefly, the sequencing data from each individual were jointly genotyped for high confidence alleles using GATK v3.8 according to the best practice. SNPs with inbreeding coefficients < −0.3, proportions of samples with high genotype-quality (GQ) (≥20) ≤ 0.8, proportions of samples with enough sequencing depth (≥10) ≤ 0.8, call rates ≤0.95 and Hardy-Weinberg Equilibrium p-value 1.0 × 10^−4^ were excluded.

The CKB is a population-based prospective cohort of 512,639 adults recruited between 2004 and 2008 from 10 geographical regions in China^32,33^. Due to limited histology information, analyses in CKB were conducted using all never-smoking lung cancer cases (71 males; 458 females) rather than LUAD-specific cases. Genotyping was performed on these cases and 47,601 never-smoking controls (4,662 male; 42,939 female) using custom Affymetrix (now ThermoFisher) Axiom® genotyping array with improved genome-wide coverage of common and low-frequency variation in Chinese populations. The sample call rate threshold was 95%, and the variant call rate threshold was 98%. Genotypes were imputed using the TOPMed reference panel. We used the imputed genotypes with imputation INFO > 0.3.

Informed consent was obtained from all participants, and the recruitment and data collection procedures were approved by the local ethics review committees of all participating research institutes.

### Construction of PRS

#### Single-ancestry PRS methods

##### PRS-25

We constructed a simple PRS using the 25 independent SNPs (PRS-25) that reached genome-wide significance (i.e., P<5×10^−8^) in the largest GWAS for LUAD in East Asian population to date (**Data S1**)^8^. The PRS-25 was calculated by summing the risk alleles weighed by their effect sizes (i.e., per allele log-odds ratio) obtained from a meta-analysis including only never-smokers. We note of the 25 SNPs previously identified, rs4268071 (MAF = 0.04) was not available in FLCCA, which we replaced with a SNP in high LD (r^2^=0.84), rs4266592.

##### PRS-CT

We generated a PRS using the CT method^34^ using PLINK 1.90^35^. This involved the clumping of SNPs based on linkage disequilibrium (LD) with a threshold of r^2^=0.1 within a 500kb window **Data S1**). The LD were estimated from half of the FLCCA dataset used for tuning dataset. We created nine SNP subsets by applying incremental P-value thresholds (i.e., P<5×10^−8^, P<5×10^−7^,…, P<5×10^−1^, 1), and calculated their respective PRSs using PLINK2^35^ command “--score cols=+scoresums,-scoreavgs no-mean-imputation”. Using the tuning dataset, the AUC was calculated for each threshold to identify the most predictive P-value cutoff.

##### LDpred2-PRS

We applied the LDpred2 method, implemented in the R package bigsnpr^36^, using a Bayesian framework to estimate SNP effect sizes through a shrinkage estimator (**Data S1**). This method leverages GWAS summary statistics, incorporating a prior for effect sizes, while also accounting for LD across SNPs. Our analysis was constrained to HapMap3 variants, and we calibrated the model using a range of hyperparameters: the proportion of causal SNPs was set across a 21-point logarithmic sequence from 10^−5^ to 1, and the per-SNP heritability was set as a fraction (0.3, 0.7, 1 or 1.4) of the total heritability estimated by LD score regression^37^ divided by the number of causal SNPs. Lastly, we used the “sparse” option to set weaker effects to zero. The optimal tuning parameters were selected based on the highest AUC achieved on the tuning dataset.

##### PRS-EUR 128

We evaluated the performance of a European-derived PRS (PRS-EUR 128)^38^, which consists of 128 variants (**Data S1**) obtained from a GWAS of predominately active smokers, on East Asian populations to assess its cross-population applicability. We note that the largest GWAS conducted in European never-smokers to date have only identified a single region (*TERT/CLPTM1L*) in which SNPs reached genome-wide significance^39^, limiting the scope of a potential targeted PRS.

#### Multi-ancestry PRSs methods

##### LDpred2 PRS + PRS-EUR 128

For the multi-ancestry PRSs, we applied the weighted-PRS approach by linearly combining the most predictive single-ancestry PRS from the East Asian population (Ldpred2 PRS) with the European population-specific PRS (PRS-EUR 128) (**Data S1**). The weights of this combined PRS were calculated by applying a logistic regression on the tuning dataset in R version 4.2.0.

##### PRS-CSx

We applied PRS-CSx, a multi-ancestry polygenic prediction method that uses a Bayesian framework with a continuous shrinkage prior to estimate SNP effect sizes from GWAS summary statistics across different populations^26^. LD reference panels for European and East Asian, provided by the PRS-CSx software, were constructed from the 1000 Genomes Project (1KG) samples ^40^. We used the default setting with the gamma-gamma prior hyperparameters (a and b) at 1 and 0.5, respectively. The shrinkage parameter *ϕ* was assessed at 10^−6^, 10^−4^, 10^−2^, and 1 within the tuning dataset to select the value with highest AUC. With the optimal *ϕ*, we calculated the PRS using posterior effect-sizes from both East Asian and European populations. Weights were then estimated to linearly combine the two PRSs on the tuning dataset (**Data S1**). The final performance was then evaluated on an East Asian independent validation dataset.

##### CT-SLEB

We used the recently developed CT-SLEB (clumping and thresholding, super learning, and empirical Bayes) method to derive a PRS using East Asian and European datasets ^24^. This method extends the standard CT method by designing a two-dimensional approach to select SNPs for East Asian PRS construction. It incorporates an empirical Bayesian (EB) framework to model genetic correlations between East Asian and EUR. Following this, a super learning (SL) model is then applied to integrate multiple PRSs, each generated using distinct p-value thresholds and clumping parameters^37^. Our implementation of CT-SLEB followed the default setting with p-value thresholds *pt* = 5×10^−8^, 5×10^−7^, …, 5×10^−1^, or 1, and genetic distances *d* = 50/*r*^2^ or 100/*r*^2^, where *r*^2^ = 0.01, 0.05, 0.1, 0.2, 0.5 *or* 0.8. The clumping process for East Asian was based on the LD reference derived from the FLCCA tuning dataset, and the European LD reference was based on the 1KG European samples.

### Relative and absolute risk calculation for PRSs

To evaluate each PRS’s performance, we standardized the scores to a unit standard deviation, based on the distribution among the control group in the East Asian validation dataset. We then quantified the association between the standardized PRSs and LUAD risk using the OR and 95% CI, via logistic regression, adjusting for age and the first 10 principal components (PCs). We evaluated the predictive performance of the different PRS models through the adjusted AUC values, accounting for age and top 10 PCs, using the R package RISCA^41^. We further estimated the ORs of each PRS for risk of LUAD based on PRS percentiles, setting the middle quantile (40-60%) as the reference category. Lastly, we used the likelihood ratio test to evaluate the interaction between the PRS and age (<40, 40-49, 50-59, 60-69, ≥70). Statistical tests performed were two-sided and an alpha<0.05 was considered noteworthy.

For absolute risk estimates, we used the iCARE software^42^ to calculate the cumulative lifetime (age 30-80) and 10-year absolute risks of LUAD among never-smoking female controls in FLCCA (N=4,544). Absolute risks were derived by applying the Cox proportional hazard model with the top performing PRS (CT-SLEB) and age-specific lung cancer incidence and mortality rates in Taiwan^43^. We chose to use incidence rates from Taiwan as approximately a quarter of FLCCA samples are from Taiwan, and most individuals in Taiwan are Han Chinese, similar to those in Mainland China. Notably, the lung cancer rates available for Taiwan are specifically for never-smokers, which makes for more appropriate estimate of absolute risks that may otherwise be inflated if ever smokers are included in the reported rates. We estimated the absolute risk of LUAD overall, as well as stratified by first-degree family history of lung cancer.

### Projections of CT and LDpred2 PRS performance by sample size

We used GENESIS package ^44^ to estimate the sample sizes needed for PRS to reach various AUC levels in the East Asian population. This method estimates the expected number of SNP discoveries and their explained heritability in future studies. Using the GWAS summary statistics from our East Asian training dataset, and the provided LD scores for East Asian populations from the 1KG dataset, we projected the AUC for CT PRS across various case-control ratios, from 1:1 to 1:10, and case numbers ranging from 5,000 to 200,000. Given GENESIS’s specific design for CT PRS, we extended its application to LDpred2 PRS projections. This involved modeling the relationship between effective sample sizes with the phenotypic variance ratio between LDpred2 and CT PRS (**Figure S1)**, elaborated in **Supplementary Methods**.

## Results

### Development and validation of the PRS

We applied several cutting-edge single-and multi-ancestry PRS methods (**Methods, Figure S2**), evaluating their performance in terms of relative risk and AUC within the FLCCA dataset of East Asian female never-smokers (**Methods, Table 1, Figure S2-S3)**. Among single-ancestry PRS methods, the LDpred2 PRS, incorporating 942,591 SNPs, outperformed other methods, with an odds ratio (OR) per unit standard deviation (SD) of 1.62 (95% confidence interval (CI): 1.52, 1.73) alongside an adjusted AUC of 0.629 (95% CI: 0.618, 0.641). In contrast, the PRS designed exclusively for European populations underperformed in our East Asian population, with an estimated adjusted AUC of 0.489 (95% CI: 0.477, 0.501), possibly attributed to the inclusion of smokers in the development of the existing European PRS.

**Table 1.**
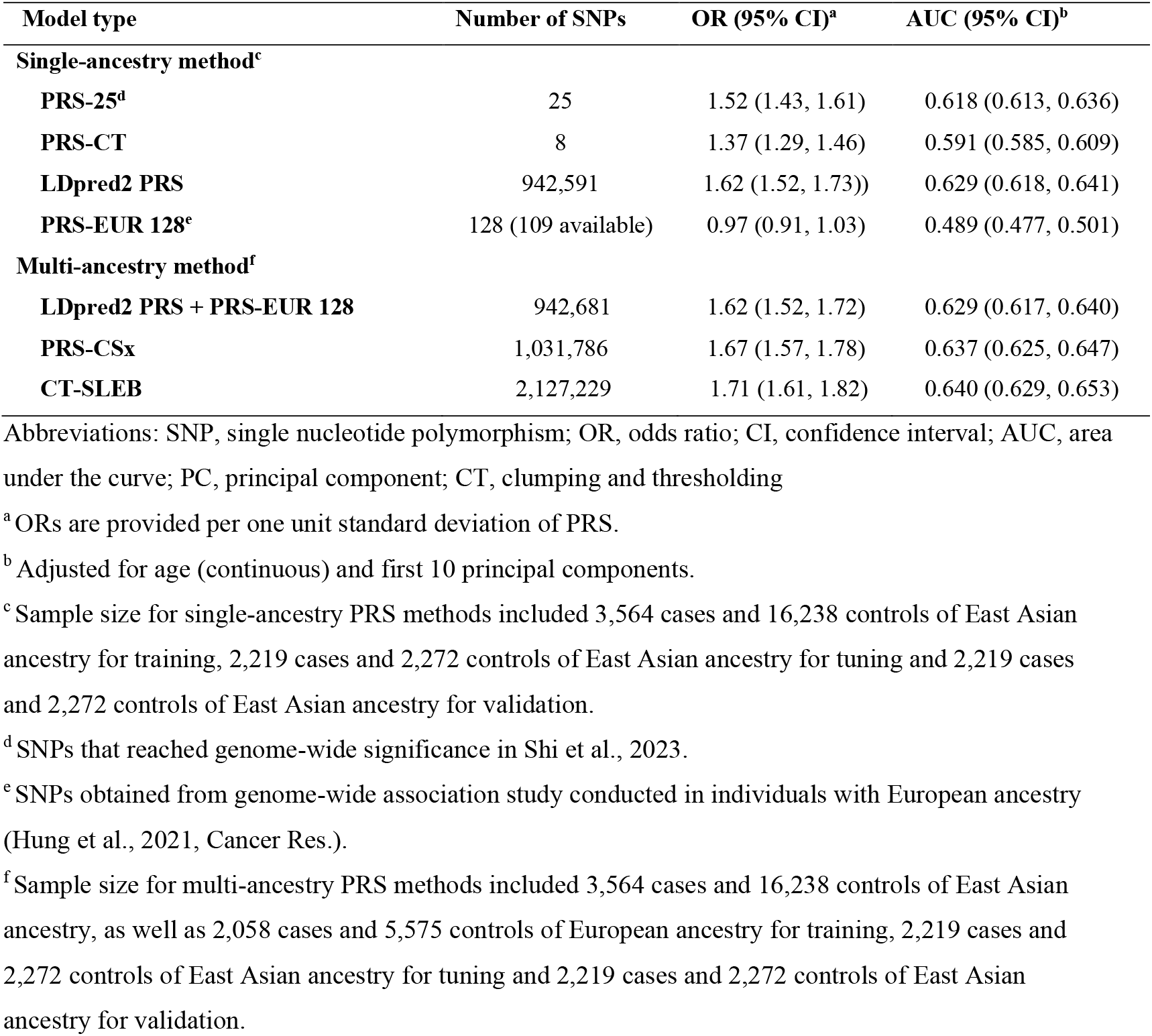
Prediction performance of different methods for generating polygenic risk scores for lung cancer in never-smoking East Asian populations in FLCCA.

| Model type | Number of SNPs | OR (95% CI)<sup>a</sup> | AUC (95% CI)<sup>b</sup> |
| --- | --- | --- | --- |
| **Single-ancestry method<sup>c</sup>** |  |  |  |
| **PRS-25<sup>d</sup>** | 25 | 1.52 (1.43, 1.61) | 0.618 (0.613, 0.636) |
| **PRS-CT** | 8 | 1.37 (1.29, 1.46) | 0.591 (0.585, 0.609) |
| **LDpred2 PRS** | 942,591 | 1.62 (1.52, 1.73) | 0.629 (0.618, 0.641) |
| **PRS-EUR 128<sup>e</sup>** | 128 (109 available) | 0.97 (0.91, 1.03) | 0.489 (0.477, 0.501) |
| **Multi-ancestry method<sup>f</sup>** |  |  |  |
| **LDpred2 PRS + PRS-EUR 128** | 942,681 | 1.62 (1.52, 1.72) | 0.629 (0.617, 0.640) |
| **PRS-CSx** | 1,031,786 | 1.67 (1.57, 1.78) | 0.637 (0.625, 0.647) |
| **CT-SLEB** | 2,127,229 | 1.71 (1.61, 1.82) | 0.640 (0.629, 0.653) |

Abbreviations: SNP, single nucleotide polymorphism; OR, odds ratio; CI, confidence interval; AUC, area under the curve; PC, principal component; CT, clumping and thresholding
<sup>a</sup> ORs are provided per one unit standard deviation of PRS.
<sup>b</sup> Adjusted for age (continuous) and first 10 principal components.
<sup>c</sup> Sample size for single-ancestry PRS methods included 3,564 cases and 16,238 controls of East Asian ancestry for training, 2,219 cases and 2,272 controls of East Asian ancestry for tuning and 2,219 cases and 2,272 controls of East Asian ancestry for validation.
<sup>d</sup> SNPs that reached genome-wide significance in Shi et al., 2023.
<sup>e</sup> SNPs obtained from genome-wide association study conducted in individuals with European ancestry (Hung et al., 2021, Cancer Res.).
<sup>f</sup> Sample size for multi-ancestry PRS methods included 3,564 cases and 16,238 controls of East Asian ancestry, as well as 2,058 cases and 5,575 controls of European ancestry for training, 2,219 cases and 2,272 controls of East Asian ancestry for tuning and 2,219 cases and 2,272 controls of East Asian ancestry for validation.

Among multi-ancestry PRS methods, the weighted-PRS of East Asian and European yielded an OR per unit SD of 1.62 (95% CI: 1.52, 1.72), with PRS-CSx and CT-SLEB showing even stronger association with ORs of 1.67 (95% CI: 1.57, 1.78) and 1.71 (95% CI: 1.61, 1.82), respectively. Among all the PRS methods, CT-SLEB had the highest AUC of 0.640 (95% CI: 0.629, 0.653). Setting individuals in the middle PRS quantile (40^th^ to 60^th^ percentile) as the reference category, those in the highest 5% of risk for the top performing PRS, CT-SLEB, had 4.17 (95% CI: 3.20, 5.47)-fold risk, whereas those in the lowest 5% had 0.33 (95% CI: 0.21, 0.51)-fold risk of developing LUAD (**Figure 2**).

**Figure 2.**
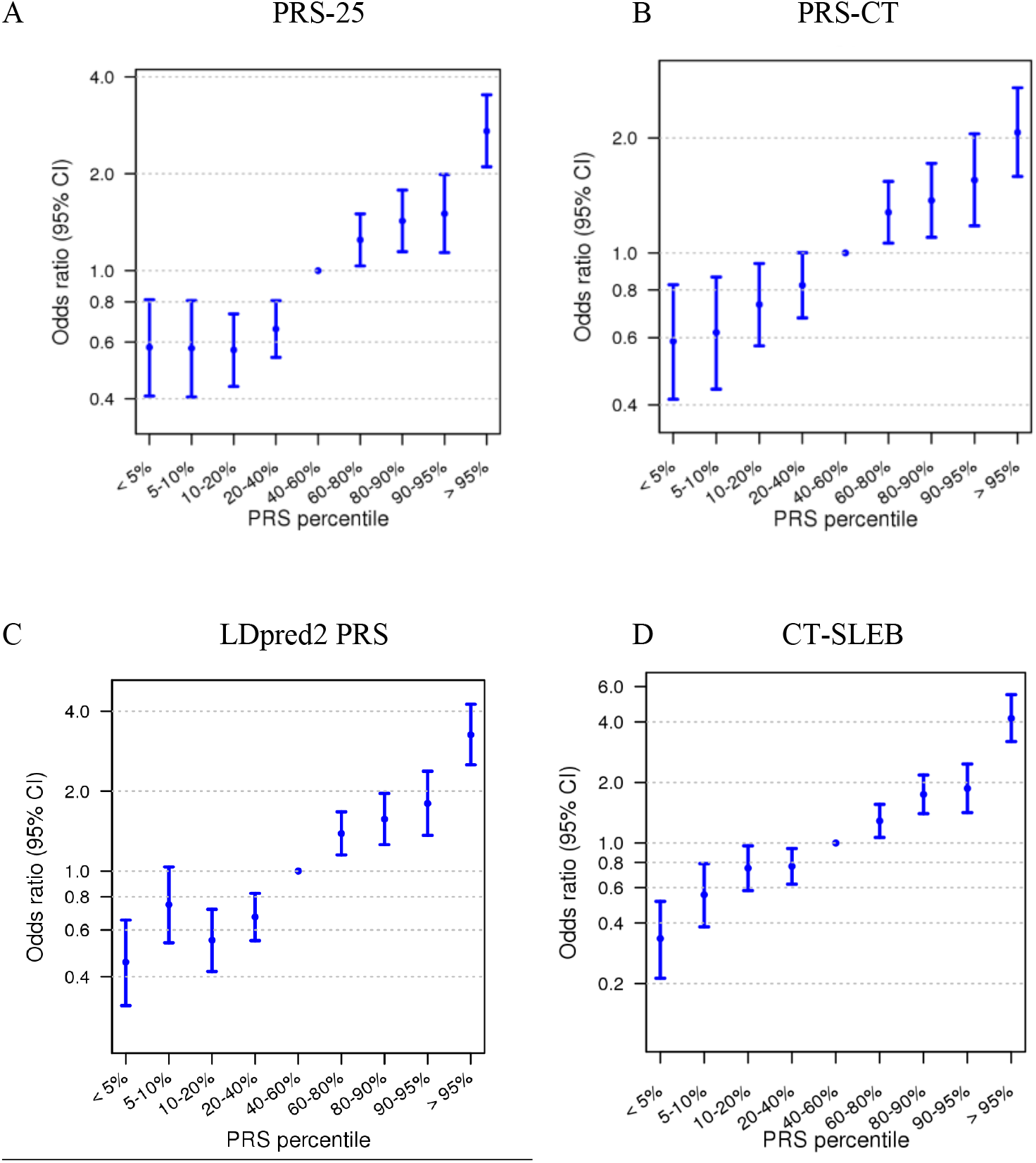
Relative risk estimated for quantiles of each polygenic risk score (PRS) and lung adenocarcinoma in the validation dataset of women with East Asian ancestry, treating the 40^th^ to 60^th^ percentile as the references. Odds ratios of PRS per standard deviation (SD) and 95% confidence intervals are shown for (A) the single-ancestry 25 SNP polygenic risk score, PRS-25 (A), (B) Clumping and thresholding method, PRS-CT, (C) Bayesian-based genome-wide approach, LDpred2 PRS, and (D) multi-ancestry approach, CT-SLEB.

We further evaluated the PRSs in independent external datasets of male and female never-smokers from NJLCC and CKB. Similar to our findings in FLCCA, we observed the LDpred2 PRS had moderate performance among the single-ancestry PRS methods with an OR of 1.77 (95% CI: 1.61, 1.95) and an adjusted AUC of 0.645 (95% CI: 0.615, 0.672) in NJLCC female never-smokers (**Table 2**). Among the multi-ancestry PRS methods, CT-SLEB yielded an OR of 1.85 (95% CI: 1.68, 2.04) and an AUC of 0.649 (95% CI: 0.623, 0.678). Notably, among male never-smokers, LDpred2 PRS yielded the highest relative risk estimate (OR=1.91, 95% CI: 1.66, 2.21) and AUC value (AUC=0.669, 95% CI: 0.633, 0.705) among both the single- and multi-ancestry PRSs. However, the estimates for the different PRS approaches were not statistically different. PRS performance in CKB was lower than in FLCCA and NJLCC, likely due to small sample size and inablility to restrict analyses to LUAD cases only. Specifically, across both the single- and multi-ancestry PRSs, PRS-25 showing the highest performance in male (OR=1.29, 95% CI: 1.02, 1.63; AUC=0.611, 95% CI: 0.547, 0.673) and female (OR=1.25, 95% CI: 1.14, 1.37); AUC=0.572, 95% CI: 0.542, 0.606) never-smokers (**Table S1)**. However, the validation was conducted among lung cancer cases with all histological subtypes, which may have limited the performance of the PRSs developed for LUAD.

**Table 2.** Prediction performance of different methods for generating polygenic risk scores for lung cancer in never-smoking East Asian populations from Nanjing, China.

| Model type | Number of SNPs | Overall |  | Male |  | Female |  |
| --- | --- | --- | --- | --- | --- | --- | --- |
|  |  | OR (95% CI)<sup>a</sup> | AUC (95% CI)<sup>b</sup> | OR (95% CI)<sup>a</sup> | AUC (95% CI)<sup>b</sup> | OR (95% CI)<sup>a</sup> | AUC (95% CI)<sup>b</sup> |
| Single-ancestry method<sup>c</sup> |  |  |  |  |  |  |  |
| PRS-25<sup>d</sup> | 25 | 1.51 (1.40, 1.63) | 0.615 (0.592, 0.638) | 1.50 (1.32, 1.73) | 0.616 (0.576, 0.654) | 1.52 (1.38, 1.67) | 0.602 (0.575, 0.632) |
| PRS-CT | 8 | 1.33 (1.23, 1.43) | 0.574 (0.549, 0.600) | 1.40 (1.23, 1.60) | 0.593 (0.553, 0.637) | 1.30 (1.19, 1.42) | 0.561 (0.529, 0.59) |
| LDpred2 PRS | 942,591 | 1.80 (1.67, 1.95) | 0.657 (0.635, 0.678) | 1.91 (1.66, 2.21) | 0.669 (0.633, 0.705) | 1.77 (1.61, 1.95) | 0.645 (0.615, 0.672) |
| PRS-EUR 128<sup>e</sup> | 128 (109 available) | 1.04 (0.96, 1.12) | 0.507 (0.481, 0.533) | 1.05 (0.92, 1.20) | 0.521 (0.473, 0.561) | 1.03 (0.94, 1.12) | 0.506 (0.473, 0.538) |
| Multi-ancestry method<sup>f</sup> |  |  |  |  |  |  |  |
| LDpred2 PRS + PRS-EUR 128 | 942,681 | 1.79 (1.65, 1.94) | 0.656 (0.632, 0.675) | 1.91 (1.66, 2.21) | 0.668 (0.631, 0.700) | 1.75 (1.59, 1.93) | 0.642 (0.617, 0.67) |
| PRS-CSx | 1,031,786 | 1.81 (1.67, 1.96) | 0.654 (0.630, 0.676) | 1.87 (1.63, 2.16) | 0.659 (0.619, 0.694) | 1.81 (1.64, 1.99) | 0.645 (0.613, 0.672) |
| CT-SLEB | 2,127,229 | 1.78 (1.64, 1.92) | 0.653 (0.631, 0.675) | 1.71 (1.49, 1.96) | 0.653 (0.618, 0.692) | 1.85 (1.68, 2.04) | 0.649 (0.623, 0.676) |

Abbreviations: SNP, single nucleotide polymorphism; OR, odds ratio; CI, confidence interval; AUC, area under the curve; PC, principal component; CT, clumping and thresholding
<sup>a</sup> ORs are provided per one unit standard deviation of PRS.
<sup>b</sup> Adjusted for age (continuous) and first 10 principal components.
<sup>c</sup> Sample size for single-ancestry PRS methods included 3,564 cases and 16,238 controls of East Asian ancestry for training, 2,219 cases and 2,272 controls of East Asian ancestry for tuning and 1,643 never-smoking LUAD cases (473 male; 1,170 female) and 2,235 never-smoking controls (795 male; 1,440 female) of East Asian ancestry for validation.
<sup>d</sup> SNPs that reached genome-wide significance in Shi et al., 2023.
<sup>e</sup> SNPs obtained from genome-wide association study conducted in individuals with European ancestry (Hung et al., 2021, Cancer Res.).
<sup>f</sup> Sample size for multi-ancestry PRS methods included 3,564 cases and 16,238 controls of East Asian ancestry, as well as 2,058 cases and 5,575 controls of European ancestry for training, 2,219 cases and 2,272 controls of East Asian ancestry for tuning and 1,643 never-smoking LUAD cases (473 male; 1,170 female) and 2,235 never-smoking controls (795 male; 1,440 female) of East Asian ancestry for validation.

### Interaction between CT-SLEB PRS and age

We observed a statistically significant multiplicative interaction between the CT-SLEB PRS and age at diagnosis for LUAD risk (p-interaction = 0.002, **Figure 3, Table S2**), with risk estimates increasing across middle-age categories (40-69) and peaking between ages 60-69 (OR = 1.93, 95% CI: 1.79-2.08). Individuals under 40 also exhibited an elevated PRS-associated risk (OR = 1.70, 95% CI: 1.42, 2.05). However, while an external validation in NJLCC was suggestive of an interaction (p-interaction = 0.07), the risk estimates followed a steady increasing trend with age, reaching the highest OR in individuals 70 and older (OR = 2.04, 95% CI: 1.64-2.58). When stratified by sex, NJLCC results were further attenuated. External validation in CKB also did not confirm an interaction between the PRS and age at diagnosis (p-interaction=0.27). While findings in FLCCA suggest a potential age-related variation in PRS effects, the lack of statistical significance in NJLCC and CKB indicates that this relationship may not be robust. Additional studies with larger sample sizes are needed to determine whether PRS effects on LUAD risk truly differ by age.

**Figure 3.**
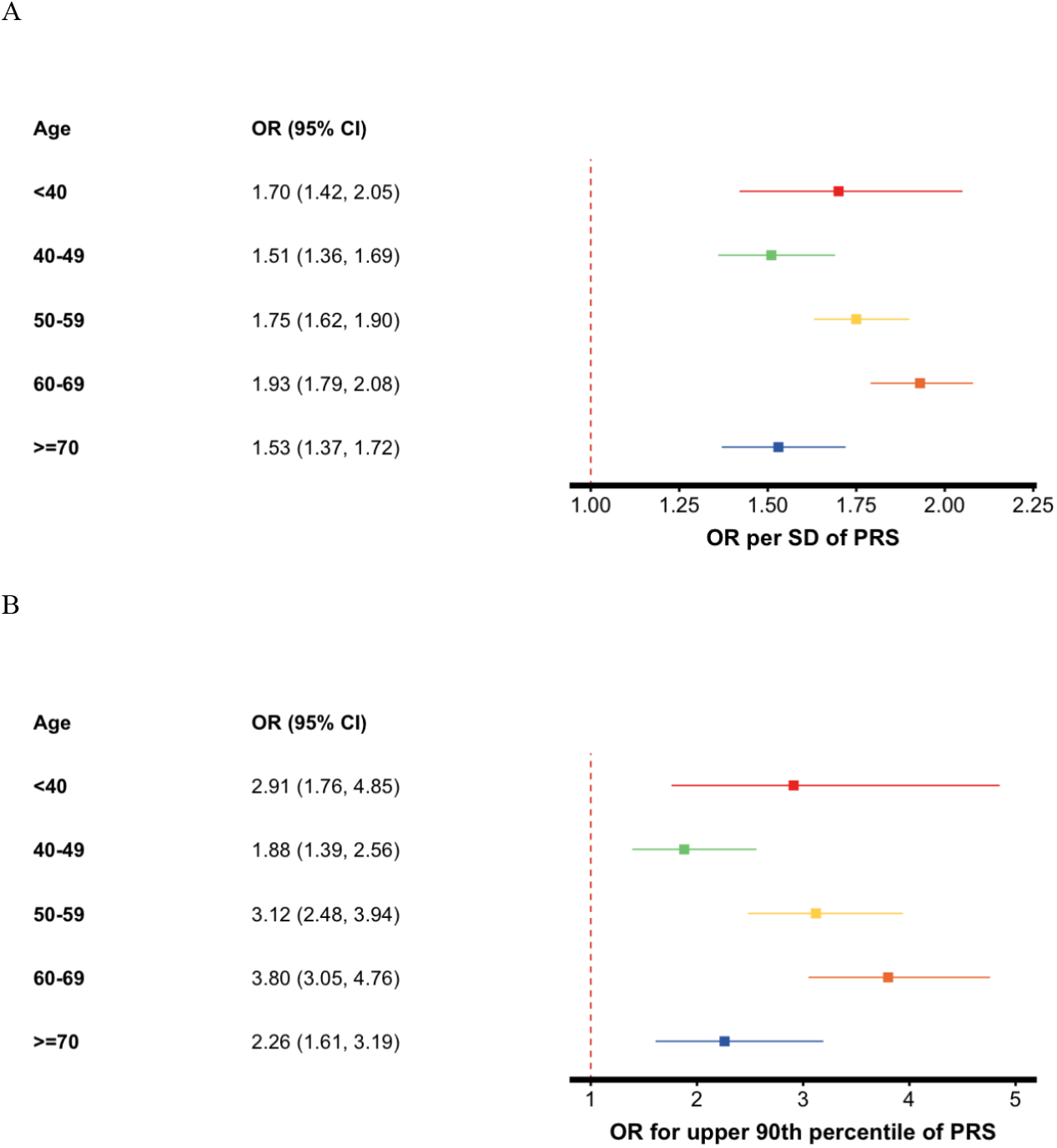
Association between polygenic risk score (PRS) and lung adenocarcinoma by age groups. (A) Odds ratios (ORs) per standard deviation (SD) of the PRS and 95% confidence intervals, and (B) ORs for individuals in the upper 90th percentile of the PRS.

### Absolute risk of developing lung adenocarcinoma by CT-SLEB PRS

Using the iCARE package^45^, we estimated the absolute risk of LUAD for never-smoking East Asian women, utilizing the CT-SLEB PRS which demonstrated the highest AUC (**Table 1**). The cumulative lifetime risk of LUAD, assessed between age 30-80, varied markedly across PRS percentiles, ranging from a minimal mean of 0.78% in the 5th percentile to a substantial mean of 6.69% in the 95th percentile (**Figure 4)**. Additionally, the 10-year absolute risk for LUAD in never-smoking 50-year-old women, varied from a mean of 0.13% in the 5th percentile to mean of 1.11% in the 95th percentile of the PRS distribution. While the average 10-year absolute risk for this age group is 0.42%, never-smoking women in the >95 and 90-95^th^ percentiles PRS reached this risk earlier, at age of 41 and 42, respectively.

**Figure 4.**
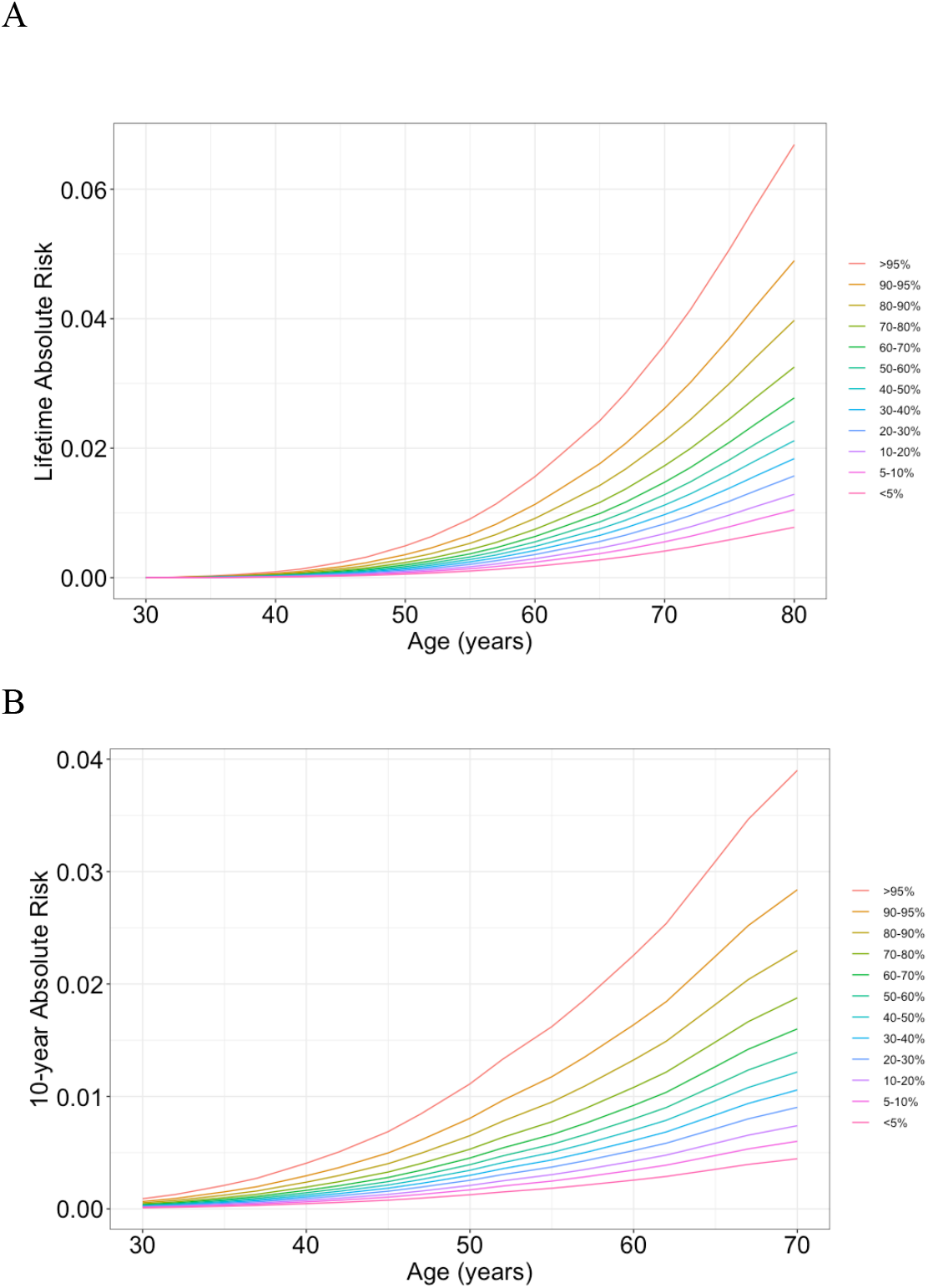
Lifetime cumulative and 10-year absolute risk of developing lung adenocarcinoma. (A) Lifetime (age 30-80) cumulative risk and (B) 10-year absolute risk of developing lung adenocarcinoma in never-smoking women in East Asia by percentiles of the CT-SLEB polygenic risk score (PRS). Absolute risks were calculated using the iCARE package^42^, based on Taiwan’s age-specific incidence and mortality data, and the PRS relative risks, as described in the Methods section.

To evaluate the combined effect of PRS and family history, we further modeled the absolute risk for LUAD in individuals with (n=86) and without (n=1,414) family history of lung cancer in a first-degree relative (**Figure S4, Figure S5**). We observed that those with family history of lung cancer have higher absolute lifetime risk of LUAD than those without family history. The lifetime absolute risk between age 30-80 for women at the bottom 5^th^ and top 95^th^ percentile of the PRS with family history of lung cancer were 2.77% and 9.39%, respectively (**Figure S4**). Additionally, the 10-year absolute risk for women in the 95^th^ PRS percentile with a family history reached the population average 10-year absolute risk at age 50 (0.42%) much earlier, at age 38. Formal tests for interaction between PRS and family history were not statistically significant (p=0.45), suggesting that family history and PRS independently contribute to LUAD risk stratification.

### Projections of polygenic risk score performance by sample size

Using the GENESIS model, we projected the expected AUC of PRS-CT and LDpred2 PRS under varying GWAS sample sizes and case-control ratios for LUAD in never smoking East Asian women (**Figure 5**). Our analysis, using the East Asian training dataset, estimated approximately 1,772 (s.e.=1,641) susceptibility variants that are independently associated with LUAD^27^. This high number of variants underscores the extensive polygenic nature of LUAD, implying relatively small effect sizes for individual SNPs. Based on our current data, the expected AUC for the LDpred2 PRS stands at 0.631 (95% CI: 0.618, 0.641), aligning closely with the actual performance of the LDpred2 PRS in the validation dataset. The upper boundary of our predictions, based solely on PRS, suggests an AUC potential of up to 0.731 (95% CI: 0.640,0.786). As sample sizes increase in our projections, the AUC is estimated to rise to 0.673 (95% CI: 0.593–0.725) with 55,000 East Asian LUAD cases and a 1:1 case-control ratio. With the same number of cases but a 1:10 case-control ratio, the AUC is projected to further increase to 0.698 (95% CI: 0.621–0.727), nearing the theoretical upper limit of predictive accuracy.

**Figure 5.**
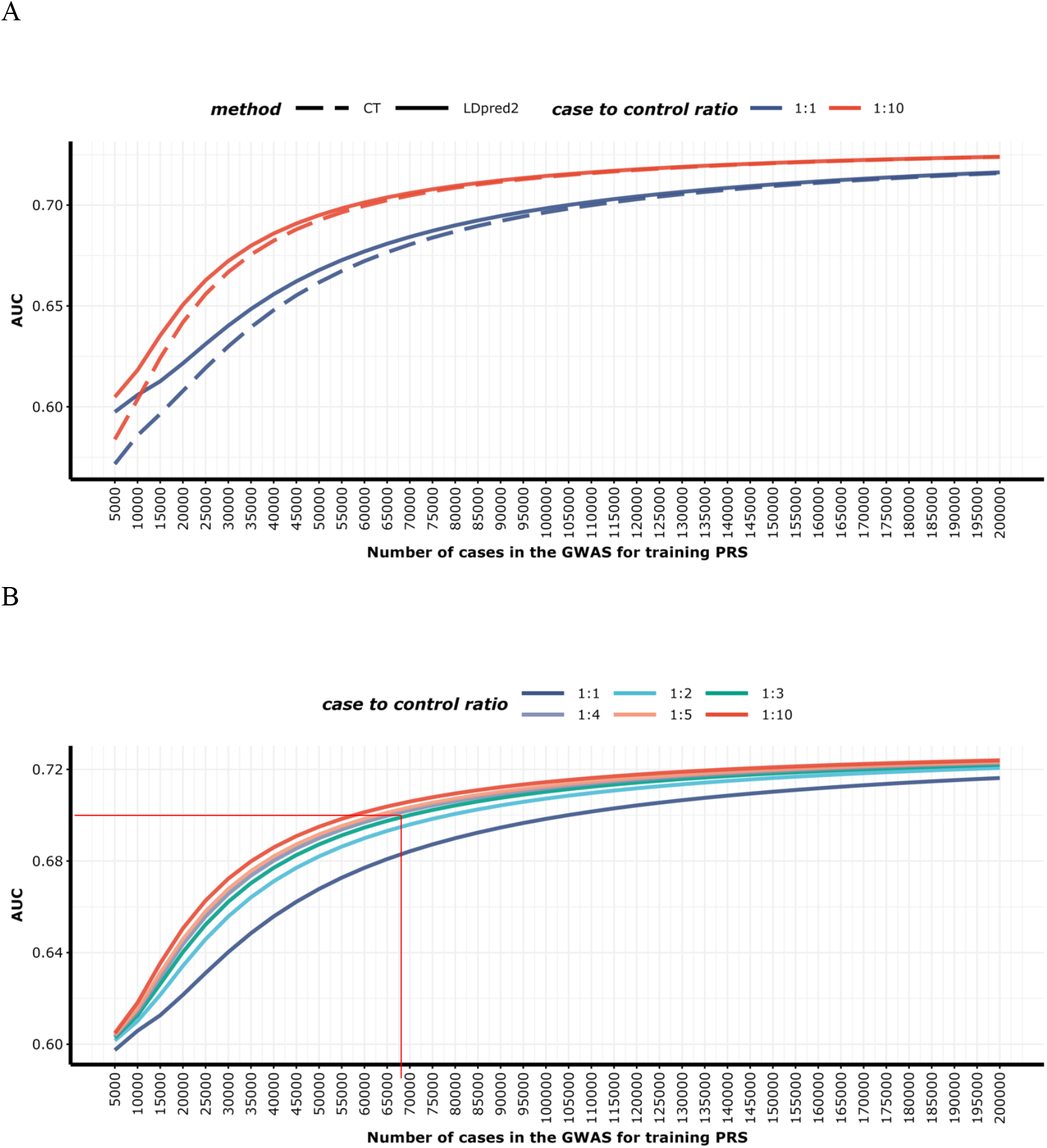
Projected area under the receiver operating characteristic curve (AUC) of polygenic risk scores (PRS) built using genome-wide association studies (GWAS) with varying sample sizes for lung adenocarcinoma in never-smoking East Asian women. (A) AUC values for PRS-CT and LDpred2 PRS with case-to-control ratios of 1:1 and 1:10. (B) AUC values for LDpred2 PRS across case-to-control ratios of 1:1, 1:2, 1:3, 1:4, 1:5, and 1:10

### Explaining genetic variance through PRS across different sample sizes

We also evaluated the genetic variance explained by the PRS under different sample sizes given the projected results (**Supplementary Methods, Table 3**). The current LDpred2 PRS explained 26.6% of the genome-wide chip heritability, contributing to approximately 16.5% of the 1.84-fold familial relative risk for lung cancer among East Asian never-smokers. Concurrently, genome-wide chip variants heritability explained 61.9% of the 1.84-fold familial relative risk associated with the disease. With an expanded sample size of 35,000 cases and 350,000 controls, the constructed PRS is projected to account for 57.9% of the genome-wide chip variants heritability, and 35.8% of the 1.84-fold familial relative risk for the disease.

**Table 3.** Genetic variance in East Asian lung cancer among never smokers explained by LDpred2 PRS.

|  | Genetic variance of PRS<sup>b</sup> | Proportion of all-GWAS variants genetic variance explained by PRS<sup>c</sup> | Proportion of Familial risk explained by PRS<sup>d</sup> |
| --- | --- | --- | --- |
| Sample size<sup>a</sup> |  |  |  |
| Current | 0.201 | 26.6% | 16.5% |
| 35,000 cases | 0.437 | 57.9% | 35.8% |
| 55,000 cases | 0.540 | 71.5% | 44.3% |

<sup>a</sup> *Sample Size*: The current sample comprises 3,564 cases and 16,238 controls. Future projections assume a 1:10 case-control ratio for sample sizes of 35,000 and 55,000 cases. Genetic variance projections for the LDpred2 PRS are based on the GENESIS method, originally designed for the CT method (refer to Nat. Genet. 50, 1318-1326 (2018)), extended to include LDpred2 by modeling the variance ratio between LDpred2 and CT (**Supplementary Methods**).
<sup>b</sup> *Genetic Variance of PRS*: This corresponds to the heritability on the frailty scale, assuming a polygenic log-additive model underpins this relationship. It quantifies the proportion of the phenotype variation that can be attributed to genetic factors in the context of PRS.
<sup>c</sup> *Proportion of Genetic Variance from All-GWAS Variants Explained by PRS*: This represents the variance of all genome-wide imputable variants as established through LD-score regression (refer to Nat. Genet. 47, 291-5 (2015) and Nat. Genet. 47, 1236-41 (2015)). On the frailty scale, the genetic variance of all GWAS variants is calculated as $\sigma_{GWAS}^2 = Var(\sum_{m=1}^M \beta_m G_m)$ , where $G_m$ is the standardized genotype for the mth SNP, $\beta_m$ is the true log odds ratio for the mth SNP and M is the total number of causal SNPs among the GWAS variants. For East Asian never smokers, the estimated genetic variance of all GWAS variants is 0.755.
<sup>d</sup> *Proportion of Familial Risk Explained by PRS*: This calculates the familial risk in terms of genetic variance using the formula $\lambda_s^2 = \exp(\sigma^2)$ , where $\lambda_s$ is the familial risk when a first-order sibling has the disease, and $\sigma^2$ is the genetic variance on frailty-scale. Further details of this calculation can be found in Nat. Genet. 31, 33-36 (2002). For lung cancer in East Asian never-smokers, the familial risk is a 1.84-fold increase, and the genetic variance of all GWAS variants, as estimated through LD-score regression, explains 61.9% of this increased familial relative risk.

## Discussion

We developed and validated single- and multi-ancestry PRSs for LUAD in never-smoking East Asian individuals using the largest GWAS dataset of never-smokers to date. The multi-ancestry PRS method, CT-SLEB, integrating summary data from East Asian and European never-smokers, emerged as the best-performing PRS among female never-smokers in FLCCA and NJLSC. It exhibited a dose-response relationship with LUAD risk and achieved higher AUC than all other evaluated PRSs. Further, our analysis demonstrates the potential of PRS in stratifying individuals’ 10-year and lifetime risk of developing LUAD. Lastly, we projected the expected discriminatory accuracy of the PRS across a range of sample sizes and case-control ratios.

To date, GWAS and subsequent PRS models have largely centered on European populations ^46,47^, rendering them less precise when applied to non-European populations and risking the exacerbation of health disparities ^48,49^. Consistent with prior research ^15^, our study observed that a lung cancer PRS constructed using European data, which included both smokers and never-smokers, significantly underperformed when applied to East Asian never-smoking individuals (AUC = 0.489, 95% CI: (0.477, 0.501)), further highlighting the heterogeneity of the disease across populations and the need to expand risk assessment efforts to non-European populations. Notably, by utilizing novel methods to integrate GWAS data from both European and East Asian populations, we demonstrated that a multi-ancestry PRS enhances the precision of risk stratification for LUAD among East Asian never-smokers.

Large-scale efforts to integrate PRS into clinical practice have focused on conditions with well-established PRS prediction performance^16–18^, such as Type 2 Diabetes, breast cancer, and cardiovascular diseases. Lung cancer has been notably absent from these efforts, primarily due to the focus on smoking as a risk factor. This has limited the exploration and application of PRS in lung cancer risk stratification, particularly among never-smokers. Recent efforts, such as the TALENT study in Taiwan^50^, have evaluated low-dose computed tomography (LDCT) screening for never-smokers with traditional risk factors like family history, passive smoking exposure, and chronic lung diseases. However, a PRS has not yet been integrated into investigated screening protocols. Further, a recent study in Taiwan incorporating genetic susceptibility in a risk model with lifestyle and environmental risk factors observed an improvement in the AUC from 0.697 to 0.714^43^. Notably, the study found that even by incorporating only 11 susceptibility loci, the model’s discriminative power had improved. Our study demonstrates the potential of a genome-wide PRS to complement these efforts, enhancing risk stratification and identifying high-risk individuals at younger ages, who may benefit from earlier interventions.

We address this gap by presenting a PRS that is associated with a 3.92-fold increase in lung cancer risk for individuals in the top 20% risk quantile, a marked improvement over the 2.09-fold increase observed in a prior study by Wei et al. ^51^ for a cohort of Chinese never-smoking women. Further, we evaluated the interaction between the PRS and age in FLCCA, NJLCC and CKB. The relationship between the PRS and age at diagnosis showed inconsistent patterns across datasets. In FLCCA, PRS effects varied nonlinearly, peaking at ages 60-69 and remaining elevated in younger individuals, suggesting a potential age-related interaction. However, NJLCC showed a gradual increase in PRS-associated risk across age groups, and estimates in CKB did not follow a specific patter. In both studies, the interaction was not statistically significant. This discrepancy may reflect differences in sample size, population structure, or environmental exposures, which could influence genetic risk expression across age groups. Future studies should examine PRS-age interactions across diverse East Asian populations, considering additional risk factors such as environmental and hormonal influences.

We estimated the lifetime and 10-year absolute risk of LUAD using the PRS, which can be used in risk stratification efforts and to identify those who are at high risk for the disease. Specifically, we found that the 10-year absolute risk for LUAD in a never-smoking 50-year-old woman, a critical age for initiating recommended annual lung cancer screening as per the United States Preventive Services Taskforce (USPSTF)^52^, varied from 0.13% in the 5th percentile to 1.11% in the 95th percentile of the PRS distribution. Notably, women in the highest risk categories (>95th and 90-95th percentiles) reached the average 10-year absolute risk (0.42%) much earlier, at ages 41 and 42, respectively. This finding suggests that the PRS has the potential to identify younger women who may benefit from earlier risk-based interventions. While limited in sample size, we also found that the lifetime absolute risk of LUAD was even higher among women in the top percentile of the PRS and with first-degree family history of lung cancer, suggesting the two risk factors may act independently for their effect on LUAD risk. Although the absolute risk values are modest, the PRS’s ability to stratify women into higher-risk categories at younger ages suggests potential utility in guiding early screening decisions. Early identification through PRS could be particularly valuable for never-smoking individuals, who are traditionally excluded from lung cancer screening programs.

The CT-SLEB method’s improved performance can be attributed to its ability to utilize diverse genotyping arrays, thereby including population-specific variants. This aspect, along with the inclusion of European samples in the training set, broadens the genetic diversity and leverages larger sample sizes for improved model accuracy. Moreover, CT-SLEB excels in modeling genetic correlations across ancestries, efficiently using both shared and unique genetic markers for refined disease risk estimations across populations. Lastly, we set up our study with a robust three-sample split design, dividing datasets into training, tuning, and validation. This ensures that PRS efficacy is validated independently, effectively reducing the overfitting risk.

Current PRS studies predominantly focus on evaluating relative and absolute risks^8,12,15,51,53^, yet our research introduces a crucial forecast of sample sizes required for differing levels of PRS accuracy. Our projection analyses indicated that achieving an AUC of 0.70 would approach the maximized prediction potential of the PRS, given the estimated genetic variance explained by GWAS chip variants. To reach this level of accuracy, a future study would need to include 55,000 cases with a 1:10 case-control ratio. Notably, accessing large biobanks with publicly available controls could reduce the number of required cases^54^. These projections offer a strategic framework for planning and designing future genetic studies on lung cancer, establishing clear benchmarks for PRS model performance, which is essential for the scientific community in advancing genetic risk prediction.

Our study has several limitations. First, it focuses solely on the genetic susceptibility of LUAD without incorporating additional questionnaire-based risk factors, especially familly history. However, leveraging the largest GWAS dataset available, our primary goal was to develop the most effective PRS model, which can be integrated with other risk indicators in future studies. Second, our projections are based on single-ancestry PRS models and do not fully account for the complexities of multi-ancestry approaches, such as varying sample sizes and genetic correlations across populations. Third, our validation cohort, derived from the FLCCA and covering various East Asian regions, relied on Taiwanese incidence data for absolute risk estimations. Lastly, while genome-wide PRS models like LDpred2 and CT-SLEB demonstrated strong performance, their reliance on nearly one million SNPs poses challenges related to overfitting and clinical feasibility. Future work should aim to optimize PRS models by prioritizing key loci while maintaining predictive power.

In summary, our study evaluates various PRS models to capture the genetic predisposition to LUAD among never-smoking East Asian individuals. It extends beyond risk prediction by estimating both 10-year and cumulative lifetime absolute risks, and by projecting the sample sizes required for future GWAS to refine the predictive power of PRSs in future GWAS. Additionally, we quantify the phenotypic variance captured by PRSs across different sample sizes. Despite advancements in using PRS for smoking cessation trials^55^, its implementation for never-smokers in clinical settings has been limited, presenting a significant area for research. Future studies are crucial to further improve these PRS models, aiming to enhance genetic risk predictions while integrating a wider array of risk factors. Such efforts will develop more accurate and comprehensive risk models for LUAD in never-smoking individuals across different populations.

## Data availability statement

All data supporting the findings described in this paper are available in the paper and in the Supplementary Information and from the corresponding author or as otherwise indicated upon request. The individual genotype data for the FLCCA data are in dbGaP phs000716.v1.p1 (Genome-Wide Association Study of Lung Cancer Susceptibility in Never-Smoking Women in Asia). For the NCC and ACC studies, please contact Kouya Shiraishi at or Takashi Kohno at for summary statistics. The GWAS data for the European populations contributing to this study are available at dbGap under accession phs000877.v1.p1 (Transdisciplinary Research Into Cancer of the Lung (TRICL), https://www.ncbi.nlm.nih.gov/projects/gap/cgi-bin/study.cgi?study_id=phs000876.v2.p1), phs001273.v3.p2 (Oncoarray Consortium, https://www.ncbi.nlm.nih.gov/projects/gap/cgi-bin/study.cgi?study_id=phs001273.v3.p2). To gain access to all data in dbGaP cited in this paper, please apply for dbGaP Authorized Access. GWAS data for the European populations contributing to this study are available under accession phs000877.v1.p1 (ILLCO), phs001273.v1.p1 (OncoArray).

## Code Availability

Methods implemented and their corresponding repositories include: SCT and LDpred2 at https://github.com/privefl/bigsnpr, PRS-CSx at https://github.com/geNan107/PRScsx. CT-SLEB at https://github.com/andrewhaoyu/CTSLEB, and GENESIS at https://github.com/yandorazhang/GENESIS. PLINK: https://www.cog-genomics.org/plink/1.9. Most of our staNsNcal analyses were performed using the following R packages: ggplot2 v.3.3.3, dplyr v.1.0.4, data.table v.1.13.6, iCARE v.1.30.0 https://github.com/KevinWFred/PRS_EASLC

## Funding

Female Lung Cancer Consortium in Asia (NCI): This study was supported by a Grant-in-Aid for Scientific Research on Priority Areas from the Ministry of Education, Science, Sports, Culture and Technology of Japan, a Grant-in-Aid for the Third Term Comprehensive 10-Year Strategy for Cancer Control from the Ministry Health, Labor and Welfare of Japan, by Health and Labor Sciences Research Grants for Research on Applying Health Technology from the Ministry of Health, Labor and Welfare of Japan, by the National Cancer Center Research and Development Fund, the National Research Foundation of Korea (NRF) grant funded by the Korea government (MEST) (grant No. 2011-0016106), a grant of the National Project for Personalized Genomic Medicine, Ministry for Health & Welfare, Republic of Korea (A111218-11-GM04), the Program for Changjiang Scholars and Innovative Research Team in University in China (IRT_14R40 to K.C.), the National Science & Technology Pillar Program (2011BAI09B00), MOE 111 Project (B13016), the National Natural Science Foundation of China (No. 30772531, and 81272618), Guangdong Provincial Key Laboratory of Lung Cancer Translational Medicine (No. 2012A061400006), Special Fund for Research in the Public Interest from the National Health and Family Planning Commission of PRC (No. 201402031), and the Ministry of Science and Technology, Taiwan (MOST 103-2325-B-400-023 & 104-2325-B-400-012). The Japan Lung Cancer Study (JLCS) was supported in part by the Practical Research for Innovative Cancer Control from Japan Agency for Medical Research and Development (15ck0106096h0002) and the Management Expenses Grants from the Government to the National Cancer Center (26-A-1) for Biobank. BioBank Japan was supported by the Ministry of Education, Culture, Sports, Sciences and Technology of the Japanese government. The Japan Public Health Center-based prospective Study (the JPHC Study) was supported by the National Cancer Center Research and Development Fund (23-A-31[toku], 26-A-2, 29-A-4, 2020-J-4, and 2023-J-4) (since 2011) and a Grant-in-Aid for Cancer Research from the Ministry of Health, Labour and Welfare of Japan (from 1989 to 2010). The Taiwan GELAC Study (Genetic Epidemiological Study for Lung AdenoCarcinoma) was sup-ported by grants from the National Research Program on Genomic Medicine in Taiwan (DOH99-TD-G-111-028), the National Research Program for Biopharmaceuticals in Taiwan (MOHW 103-TDUPB-211-144003, MOST 103-2325-B-400-023) and the Bioinformatics Core Facility for Translational Medicine and Biotechnology Development (MOST 104-2319-B-400-002). This work was also supported by the Jinan Science Research Project Foundation (201102051), the National Key Scientific and Technological Project (2011ZX09307-001-04), the National Natural Science Foundation of China (No.81272293), the State Key Program of National Natural Science of China (81230067), the National Research Foundation of Korea (NRF) grant funded by the Korea government (MSIP) (No. NRF-2014R1A2A2A05003665), Sookmyung Women’s University Research Grants, Korea (1-1603-2048), Agency for Science, Technology and Research (A*STAR), Singapore and the US National Institute of Health Grant (1U19CA148127-01). The overall GWAS project was supported by the intramural program of the US National Institutes of Health/National Cancer Institute. The following is a list of grants by study center: SKLCS (Y.T.K.)—National Research Foundation of Korea (NRF) grant funded by the Korea government (MEST) (2011-0016106). (J.C.) – This work was supported by a grant from the National R&D Program for Cancer Control, Ministry of Health &Welfare, Republic of Korea (grant no. 0720550-2). (J.S.S) – grant number is A010250. WLCS (T.W.)—National Key Basic Research and Development Program (2011CB503800). SLCS (B.Z.)—National Nature Science Foundation of China (81102194). Liaoning Provincial Department of Education (LS2010168). China Medical Board (00726). GDS (Y.L.W.)—Foundation of Guangdong Science and Technology Department (2006B60101010, 2007A032000002, 2011A030400010). Guangzhou Science and Information Technology Bureau (2011Y2-00014). Chinese Lung Cancer Research Foundation, National Natural Science Foundation of China (81101549). Natural Science Foundation of Guangdong Province (S2011010000792). TLCS (K.C., B.Q)—Program for Changjiang Scholars and Innovative Research Team in University (PCSIRT), China (IRT1076). Tianjin Cancer Institute and Hospital. National Foundation for Cancer Research (US). FLCS (J.C.W., D.R., L.J.)—Ministry of Health (201002007). Ministry of Science and Technology (2011BAI09B00). National S&T Major Special Project (2011ZX09102-010-01). China National High-Tech Research and Development Program (2012AA02A517, 2012AA02A518). National Science Foundation of China (30890034). National Basic Research Program (2012CB944600). Scientific and Technological Support Plans from Jiangsu Province (BE2010715). NLCS (H.S.)—China National High-Tech Research and Development Program Grant (2009AA022705). Priority Academic Program Development of Jiangsu Higher Education Institution. National Key Basic Research Program Grant (2011CB503805). GEL-S (A.S.)—National Medical Research Council Singapore grant (NMRC/0897/2004, NMRC/1075/2006). (J.Liu)—Agency for Science, Technology and Research (A*STAR) of Singapore. GELAC (C.A.H.)—National Research Program on Genomic Medicine in Taiwan (DOH98-TDG-111-015). National Research Program for Biopharmaceuticals in Taiwan (DOH 100-TD-PB-111-TM013). National Science Council, Taiwan (NSC 100-2319-B-400-001). YLCS (Q.L.)—Supported by the intramural program of U.S. National Institutes of Health, National Cancer Institute. SWHS (W.Z., W-H.C., N.R.)—The work was supported by a grant from the National Institutes of Health (R37 CA70867, UM1 CA182910) and the National Cancer Institute intramural research program, including NCI Intramural Research Program contract (N02 CP1101066). JLCS (K.M., T.K.)—Grants-in-Aid from the Ministry of Health, Labor, and Welfare for Research on Applying Health Technology and for the 3rd-term Comprehensive 10-year Strategy for Cancer Control; by the National Cancer Center Research and Development Fund; by Grant-in-Aid for Scientific Research on Priority Areas and on Innovative Area from the Ministry of Education, Science, Sports, Culture and — Technology of Japan. (W.P.)—NCI R01-CA121210. HKS (J.W.)— General Research Fund of Research Grant Council, Hong Kong (781511M). The Environment and Genetics in Lung Cancer Etiology (EAGLE), Prostate, Lung, Colon, Ovary Screening Trial (PLCO), and Alpha-Tocopherol, Beta-Carotene Cancer Prevention (ATBC) studies were supported by the Intramural Research Program of the National Institutes of Health, National Cancer Institute (NCI), Division of Cancer Epidemiology and Genetics. ATBC was also supported by U.S. Public Health Service contracts (N01-CN-45165, N01-RC-45035, and N01-RC-37004) from the NCI. PLCO was also supported by individual contracts from the NCI to the University of Colorado Denver (NO1-CN-25514), Georgetown University (NO1-CN-25522), the Pacific Health Research Institute (NO1-CN-25515), the Henry Ford Health System (NO1-CN-25512), the University of Minnesota, (NO1-CN-25513), Washington University (NO1-CN-25516), the University of Pittsburgh (NO1-CN-25511), the University of Utah (NO1-CN-25524), the Marshfield Clinic Research Foundation (NO1-CN-25518), the University of Alabama at Birmingham (NO1-CN-75022), Westat, Inc. (NO1-CN-25476), and the University of California, Los Angeles (NO1-CN-25404). The Carotene and Retinol Efficacy Trial (CARET) is funded by the National Cancer Institute, National Institutes of Health through grants U01-CA063673, UM1-CA167462, and U01-CA167462. The Cancer Prevention Study-II (CPS-II) Nutrition Cohort was supported by the American Cancer Society. The NIH Genes, Environment and Health Initiative (GEI) partly funded DNA extraction and statis- tical analyses (HG-06-033-NCI-01 and RO1HL091172-01), genotyping at the Johns Hopkins University Center for Inherited Disease Research. This research was supported by the National Research Foundation of Korea (NRF) grant funded by the Korea government (MSIT) (No. 2020R1A2C4002236)

Female Lung Cancer Consortium in Asia (Tianjin): Tianjin Science and Technology Committee Foundation, 18YFZCSY00520.

Female Lung Cancer Consortium in Asia (Taiwan): The Ministry of Health and Welfare grants DOH97-TD-G-111-028 (ISC), DOH98-TD-G-111-017 (ISC), DOH99-TD-G-111-014 (ISC); DOH97-TD-G-111-026 (CAH), DOH98-TD-G-111-015 (CAH), DOH99-TD-G-111-028 (CAH); National Health Research Institutes grants NHRI-PH-110-GP-01, NHRI-PH-110-GP-03; and the Ministry of Science and Technology grants MOST108-2314-B-400-038(CAH), MOST109-2740-B-400-002(CAH), MOST 111-2740-B-400-002 (CAH), MOST 111-2314-B-400-020 (CAH).

The GWAS of lung cancer in European never smokers was supported by NIH R01 CA149462 (OYG).

OncoArray study in Europeans: The OncoArray data and analysis from INTEGRAL-ILCCO were supported by NIH U19 CA203654, and U19 CA148127. The data harmonization for ILCCO was supported by Canadian Institute for Health Research (CIHR) Canada Research Chair to R.J.H, and CIHR FDN 167273).

European never-smoking lung cancer study: CIA is a Research Scholar of the Cancer Prevention Institute of Texas (CPRIT) and supported by CPRIT grant RR170048.

Taiwan eQTL study: This study was supported by the Ministry of Health and Welfare grants DOH97-TD-G-111-028 (ISC), DOH98-TD-G-111-017 (ISC), DOH99-TD-G-111-014 (ISC); DOH97-TD-G-111-026 (CAH), DOH98-TD-G-111-015 (CAH), DOH99-TD-G-111-028 (CAH); National Health Research Institutes grants NHRI-PH-110-GP-01, NHRI-PH-110-GP-03; and the Ministry of Science and Technology grants MOST108-2314-B-400-038(CAH), MOST109-2740-B-400-002(CAH).

This research was also supported by the National Natural Science Foundation of China (82388102)

B.B., K.W., J.C., J.S. N.R., Q.L. and H.Z. are supported by NIH intramural Research Program. N.C. is supported by NIH grant 1R01HG010480. P.Y. is supported by Mayo Clinic Foundation Research Funds, NIH-CA77118 and CA80127. G.L. is supported is supported by the Alan Brown Chair and Lusi Wong Fund of the Princess Margaret Cancer Foundation. D.C.C is supported by U01CA209414. O.Y.G. is supported by NIH R01 CA231141.

This research was supported [in part] by the Intramural Research Program of the National Institutes of Health (NIH). The contributions of the NIH authors were made as part of their official duties as NIH federal employees, are in compliance with agency policy requirements, and are considered works of the United States Government. However, the findings and conclusions presented in this paper are those of the authors and do not necessarily reflect the views of the NIH or the U.S. Department of Health and Human Services.

## Abbreviations

LUAD: lung adenocarcinoma
GWAS: genome-wide association study
EUR: European
PRS: polygenic risk score
SNP: single nucleotide polymorphism
FLCCA: Female Lung Cancer Consortium in Asia
LD: linkage disequilibrium
AUC: area under the receiver operating curve
PC: principal component
OR: odds ratio
CI: confidence interval
USPSTF: United States Preventive Services Taskforce

## Conflict of Interest

The authors declare no competing interests.

## Acknowledgements

We gratefully acknowledge the important historical contribution that the following individuals made to the development and success of this long-term collaboration:

Minsun Song, PhD, Department of Statistics & Research Institute of Natural Sciences, Sookmyung Women’s University, Seoul, Republic of Korea;

Michiaki Kubo, PhD, Laboratory for Genotyping Development, RIKEN Center for Integrative Medical Sciences, Yokohama,, Japan;

Haruhiko Nakayama, PhD, Department of Thoracic Surgery, Kanagawa Cancer Center, Yokohama, Japan;

Yuichiro Ohe, PhD,Department of Thoracic Surgery, National Cancer Center Hospital, Tokyo, Japan;

Jian Su, PhD, Guangdong Lung Cancer Institute, Guangdong Provincial Key Laboratory of Translational Medicine in Lung Cancer, Guangdong Provincial People’s Hospital (Guangdong Academy of Medical Sciences), Southern Medical University, Guangzhou, China;

Ying-Huang Tsai, PhD, Department of Respiratory Therapy, Chang Gung University, Taoyuan, Taiwan;

Ming-Shyan Huang, PhD, Department of Internal Medicine, E-Da Cancer Hospital, I-Shou University and Kaohsiung Medical University, Kaohsiung, Taiwan;

Kun-Chieh Chen, PhD, Department of Internal Medicine, Division of Pulmonary Medicine, Chung Shan Medical University Hospital, Taichung, Taiwan;

Biyun Qian, PhD, Department of Epidemiology and Biostatistics, Key Laboratory of Prevention and Control of Human Major Diseases of the Ministry of Education, Key Laboratory of Molecular Cancer Epidemiology of Tianjin, National Clinical Research Center for Cancer, Tianjin Medical University Cancer Institute and Hospital, Tianjin Medical University, Tianjin, China;

Chen Wu, PhD, Department of Etiology & Carcinogenesis and State Key Laboratory of Molecular Oncology, Cancer Institute and Hospital, Chinese Academy of Medical Sciences and Peking Union Medical College, Beijing, China;

Margaret R Spitz, PhD, Department of Medicine, Section of Epidemiology and Population Science, Institute for Clinical and Translational Research, Houston, TX, USA;

Mikael Johansson, PhD, Department of Radiation Sciences, Umeå University, Umeå, Sweden;

Jae Sook Sung, PhD, Department of Internal Medicine, Division of Oncology/Hematology, College of Medicine, Korea University Anam Hospital, Seoul, Republic of Korea;

Yoo Jin Jung, PhD, Department of Thoracic and Cardiovascular Surgery, Cancer Research Institute, Seoul National University College of Medicine, Seoul, Republic of Korea;

Huan Guo, PhD, Department of Occupational and Environmental Health and Ministry of Education Key Lab for Environment and Health, School of Public Health, Tongji Medical College, Huazhong University of Science and Technology, Wuhan, China;

Bin Zhu, PhD, Division of Cancer Epidemiology and Genetics, National Cancer Institute, Rockville, MD, USA;

Li Liu, PhD, Department of Oncology, Cancer Center, Union Hospital, Huazhong University of Science and Technology, Wuhan, China;

Ying Chen, PhD, Saw Swee Hock School of Public Health, National University of Singapore and National University Health System, Singapore, Singapore;

Jun Suk Kim, PhD, Department of Internal Medicine, Division of Medical Oncology, College of Medicine, Korea University Guro Hospital, Seoul, Republic of Korea;

Ho-Il Yoon, PhD,Department of Internal Medicine, Seoul National University Bundang Hospital, Seongnam, Republic of Korea;

Ping Xu, PhD,Department of Oncology, Wuhan Iron and Steel (Group) Corporation Staff-Worker Hospital, Wuhan, China;

Chih-Liang Wang, PhD, Department of Thoracic Medicine, Division of Pulmonary Oncology and Interventional Bronchoscopy, Chang Gung Memorial Hospital, Taoyuan, Taiwan;

Wei-Yen Lim, PhD, Saw Swee Hock School of Public Health, National University of Singapore and National University Health System, Singapore, Singapore;

Hongyan Chen, PhD, Ministry of Education Key Laboratory of Contemporary Anthropology, School of Life Sciences, Fudan University, Shanghai, China;

Li Jin, PhD, Ministry of Education Key Laboratory of Contemporary Anthropology, School of Life Sciences, Fudan University, Shanghai, China;

Shengchao A. Li, MS, Division of Cancer Epidemiology and Genetics, National Cancer Institute,, Rockville, MD, USA;

Gening Jiang, PhD, Shanghai Pulmonary Hospital, Shanghai, China; Ke Fei, PhD, Shanghai Pulmonary Hospital, Shanghai,, China;

This work utilized the computational resources of the high-performance computation Biowulf cluster at National Institutes of Health, USA (http://hpc.nih.gov).

## Author contributions

B.B., N.R., Q.L. and H.Z. organized and designed the study. B.B., X.W., H.Z. carried out the statistical analyses with supervision from N.R., Q.L. and H.Z. C.K. conducted the validation analyses in CKB. J.D., Y.W., T. Q. conducted the validation analyses in NJLCC. B.B., X.W., Q.L. N.R. and H.Z. wrote the first draft. B.B., X.W., C.K., J.D. Q.L. N.R. and H.Z. review and edit the draft. Q.L., M.T.L., B.A.B., W.H., N.E.C., BT.J., M.Song, H.P., D.A., C.C.C., L.B., M.Y., A.H., B.H., J.Liu, B.Zhu, S.I.B., C.K., K.Wyatt, S.A.L., A.Chao, J.F.F.J., S.J.C., N.R., Z.Wang, C..L., J.C., C.W., W.T., D.Lin, SJ.A., XC.Z., J.S., YL.W., M.P.W., L.P.C., J.C.M.H., V.H.F.L., Z.H., K.M., J.Y..P., Jia.Liu, HS.J., J.E..C., Y.Y.C., H.N.K., MH.S., SS.K., YC.K., IJ.O., S.W.S., HI.Y., Y.T.K., YC.H., J.H.K., Y.H.K., J.S..S., Y.J..J., K.H.P., C.H.K., J.S.K., I.K.P., B..S., Jie.L, Z.W., S.C., J.Y., JC.W., Y.Y., YB.X., YT.G., D.L., J.Y.Y.W., H.C., L..J., J.Z., G.J., K..F., Z.Y., B.Z., W.W., P.G., Q.H., X.L., Y.R., A.S., Y.L., Y.C., WY.L., W.Z., XO.S., Q.C., G.Y., B.Q., T.W., H.G., L..L., P.X., F.W., G.W., J.X., J.L., R.CH.V., B.B., H.D.III.H., J.Wang, A.D.L..S., J.KC.C., V.L.S., K.C., H.Z., H.D., C.A.H., TY.C., LH.C…, IS.C., CY.C., S.S.J ., CH.C., GC.C., CF.H., YH.T., WC.W., KY.C., MS.H., WC.S., YM.C., CL.W., KC.C., CJ.Y., HH.H., FY.T.., HC.L., CJ.C., PC.Y., K.Shiraishi, T.K., H.K., S.M., H.H., K.Goto, Y.Ohe, S.W., Y.Yatabe, M.T., R.Hamamoto, A.Takahashi, Y.Momozawa, M.Kubo, Y.K., Y.D., Y.Miyagi, H.N., T.Y., N.S., M.I., M.H., Y.N., K.Takeuchi, K.W., K.Matsuda, Y.Murakami, K.S., K.T., Y.O., M.S., H.Suzuki, A.G., Y.M., T.H., M.K., K.O., H.S., J.D., H.M., M.Z., R.J.H., S.L., A.T., C.C., S.E.B., M.Johansson, A.R., H.Bö., HE.W., D.C., G.R., S.A., P.B., J.MK., J.K.F., S.S.S., L.L.M., O.M.., H.Bö., G.L., A.A., L.A.K., S.ZN., K.G., M.J., A.C., JM.Y., P.L., M.B.S., M.C.A., C.I.A., A.G.S., R.H., M.R.S., O.Y.G., I.P.G., X.W., P.Y. conducted epidemiology studies and contributed samples to GWAS and/or conducted initial genotyping. All authors reviewed and approved the final review of the manuscript.

## Notes

### Competing Interest Statement

The authors have declared no competing interest.

### Funding Statement

Details of the funding statement are provided in the acknowledgment section of the manuscript

### Author Declarations

Ethics committee/IRB of the National Cancer Center Institutional gave ethical approval for this work Ethics committee/IRB of Japan and the Aichi Cancer Center Ethics Committee, Japan gave ethical approval for this work

### Summary of Updates

We have validated the PRSs using another independent cohort study to show the portability of the results

