## Supplementary Figures and Notes for "Stratifying Lung Adenocarcinoma Risk with Multi-ancestry Polygenic Risk Scores in East Asian Never-Smokers"

Supplementary Material

### Supplementary Methods

*Model fitting for ratios of PRSs between LDpred2 and PRS-CT*

In the context of GENESIS, which is intended for sample size projections via the CT method, we established a model to describe the relationship between the effective sample size (N_eff) and the phenotypic variance ratios elucidated by CT and LDpred2 PRS. The effective sample size was denoted as $N_{eff} = N_{case} * N_{control}/(N_{case} + N_{control})$, where $N_{case}$ and $N_{control}$ represent the counts of cases and controls, respectively. Our training dataset was systematically downsized to seven distinct sample sizes, derived through combinations of the three East Asian studies included in our training data, enabling us to evaluate the performance of PRS-CT and LDpred2 PRS at each data point. This analysis yielded seven data points depicting the variance ratio between LDpred2 PRS and PRS-CT across a spectrum of sample sizes. For accurate characterization, we sought models satisfying two conditions: firstly, as $N_{eff}$ increases, the ratio of phenotypic variance explained by the two methods will converge to 1, given that both PRSs approach the heritability of the genetic effects. Secondly, this ratio should inversely correlate with the increase in sample size. Additionally, we assumed that beyond an $N_{eff}$ of 50,000, the variance ratio between PRS-CT and LDpred2 PRS would converge to 1.

We examined five mathematical models to encapsulate this relationship: exponential decay, power law, logistic, Gompertz, and Weibull functions. Defining $y$ as the ratio of phenotypic variance between LDpred2 and CT PRS, and $n$ as the East Asian population's effective sample size, the models are formulated as follows:

Exponential decay:

$$y = ae^{-bn} + c,$$

Power law:

$$y = an^{-b} + 1,$$

Logistic:

$$y = \frac{a}{1 + e^{-b(n-c)}} + (1 - a),$$

Geompertz:

$$y = \frac{a}{e^{-be^{n-c}}} + 1,$$

Weibull:

$$y = ae^{-bn^c} + 1,$$

Here, a, b, and c represent the parameters to estimate for each model. To determine the best fit, we calculated the coefficient of determination ($R^2$) for each model:

$$R^2 = 1 - \frac{\sum_i (y_i - \hat{y}_i)^2}{\sum_i (y_i - \overline{y})^2},$$

Here $\overline{y}$ is the mean of the observed values $y_i$. A model with a higher $R^2$ indicates a more accurate representation of the data.

Each model was fitted to the data points using nonlinear least squares regression, utilizing the Levenberg-Marquardt algorithm as implemented in the 'minpack.lm' package[1] within R. We selected initial parameter estimates based on the observed range of effective sample sizes and phenotypic variance ratios. The performance of each model was gauged by its R-squared value—the proportion of variance in the observed data that is predictable from the independent variables.

Of the models tested, The Weibull model was identified as the best fit (**Supplementary Figure 1**). The finalized Weibull model was:

$$y = 3.511e^{-0.049n^{0.429}} + 1.$$

This model was used to extrapolate the phenotypic variance of CT PRS to predict the variance for LDpred2. Finally, the projected phenotypic variance was translated into AUC values[2] to estimate the AUC for LDpred2 PRS at different sample sizes.

*Proportion of genetic variance explained by LDpred2 PRS across different sample sizes*

As described in the last section, we estimated the genetic variance explained by LDpred2 PRS ($\sigma^2_{LDpred2}$) under different sample sizes. This genetic variance is equated with heritability on a frailty scale, premised on the polygenic log-additive model as the fundamental genetic architecture. Specifically, the genetic variance on this scale for all GWAS variants is formulated as $\sigma^2_{GWAS} = var(\sum_{m=1}^{M} \beta_m G_m)$, where $G_m$ is the standardized genotype for the mth SNP, $\beta_m$ is the true log OR for the mth SNP and M is the total number of causal SNPs within the GWAS variants. To estimate the frailty scale heritability for lung cancer in East Asian never-smokers, we used the linkage-disequilibrium score regression[3] to estimate $\sigma^2_{GWAS}$ using summary statistics from the training dataset along with the provided East Asian -specific LDscore derived from the 1000 Genomes Project data. Consequently, the fraction of genetic variance attributable to all GWAS variants that is explained by the LDpred2 PRS is denoted by the ratio $\sigma^2_{LDpred2}/\sigma^2_{GWAS}$.

The conversion of familial risk to genetic variance employed the expression $\lambda_s^2 = exp(\sigma^2)$, where $\lambda_s$ is the familial risk when a first-order sibling has the disease, and $\sigma^2$ is the genetic variance on frailty-scale[4]. The familial risk of lung cancer in East Asian never-smokers was reported as a 1.84-fold increase[5], correspond to a genetic variance $\sigma^2$ of 1.22. Thus, the LDpred2 PRS's elucidation of familial risk is quantified by the ratio $\sigma^2_{LDpred2}/\sigma^2$.

Table S1. Prediction performance of different methods for generating polygenic risk scores for lung cancer in never-smoking East Asian populations from the China Kadoorie Biobank.

| | | Overall | | Male | | Female | |
|---|---|---|---|---|---|---|---|
| **Model type** | **Number of SNPs** | **OR (95% CI)[a]** | **AUC (95% CI)[b]** | **OR (95% CI)[a]** | **AUC (95% CI)[b]** | **OR (95% CI)[a]** | **AUC (95% CI)[b]** |
| **Single-ancestry method[c]** | | | | | | | |
| **PRS-25[d]** | 25 | 1.26 (1.15, 1.37) | 0.574 (0.544, 0.603) | 1.29 (1.02, 1.63) | 0.611 (0.547, 0.673) | 1.25 (1.14, 1.37) | 0.572 (0.542, 0.606) |
| **PRS-CT** | 8 | 1.20 (1.11, 1.31) | 0.556 (0.521, 0.588) | 1.18 (0.93, 1.49) | 0.591 (0.508, 0.660) | 1.21 (1.10, 1.32) | 0.552 (0.516, 0.590) |
| **LDpred2 PRS** | 942,591 | 1.32 (1.21, 1.44) | 0.573 (0.542, 0.604) | 1.36 (1.07, 1.73) | 0.572 (0.476, 0.655) | 1.31 (1.19, 1.44) | 0.570 (0.537, 0.602) |
| **PRS-EUR 128[e]** | 128 (109 available) | 0.98 (0.90, 1.07) | 0.490 (0.460, 0.521) | 1.02 (0.80, 1.29) | 0.515 (0.415, 0.599) | 0.98 (0.89, 1.08) | 0.484 (0.448, 0.515) |
| **Multi-ancestry method[f]** | | | | | | | |
| **LDpred2 PRS + PRS-EUR 128** | 942,681 | 1.31 (1.20, 1.43) | 0.574 (0.542, 0.602) | 1.35 (1.06, 1.71) | 0.572 (0.486, 0.653) | 1.31 (1.19, 1.43) | 0.571 (0.535, 0.605) |
| **PRS-CSx** | 1,031,786 | 1.29 (1.18, 1.40) | 0.569 (0.532, 0.605) | 1.32 (1.04, 1.70) | 0.587 (0.504, 0.671) | 1.29 (1.17, 1.41) | 0.563 (0.527, 0.600) |
| **CT-SLEB** | 2,127,229 | 1.29 (1.18, 1.41) | 0.571 (0.536, 0.605) | 1.35 (1.06, 1.72) | 0.591 (0.511, 0.669) | 1.28 (.1.17, 1.41) | 0.568 (0.528, 0.607) |

Abbreviations: SNP, single nucleotide polymorphism; OR, odds ratio; CI, confidence interval; AUC, area under the curve; PC, principal component; CT, clumping and thresholding

[a] ORs are provided per one unit standard deviation of PRS.

[b] Adjusted for age (continuous) and first 10 principal components.

[c] Sample size for single-ancestry PRS methods included 3,564 cases and 16,238 controls of East Asian ancestry for training, 529 never-smoking lung cancer cases (71 males; 458 females) and 47,601 never-smoking controls (4,662 male; 42,939 female) of East Asian ancestry for tuning and 2,219 cases and 2,272 controls of East Asian ancestry for validation.

[d] SNPs that reached genome-wide significance in Shi et al., 2023.

[e] SNPs obtained from genome-wide association study conducted in individuals with European ancestry (Hung et al., 2021, Cancer Res.).

[f] Sample size for multi-ancestry PRS methods included 3,564 cases and 16,238 controls of East Asian ancestry, as well as 2,058 cases and 5,575 controls of European ancestry for training, 2,219 cases and 2,272 controls of East Asian ancestry for tuning and 529 never-smoking lung cancer cases (71 males; 458 females) and 47,601 never-smoking controls (4,662 male; 42,939 female) of East Asian ancestry for validation.

Table S2. Association between polygenic risk scores and lung adenocarcinoma by age category in the Female Lung Cancer Consortium in Asia

| | | OR (95% CI) | P-interaction* |
|---|---|---|---|
| PRS-25 | <40 | 1.51 (1.27, 1.81) | $2.03x10^{-83}$ |
| | 40-49 | 1.27 (1.36, 1.70) | |
| | 50-59 | 1.36 (1.45, 1.70) | |
| | 60-69 | 1.55 (1.43, 1.67) | |
| | ≥70 | 1.38 (1.24, 1.54) | |
| PRS-CT | <40 | 1.59 (1.33, 1.91) | $5.50x10^{-73}$ |
| | 40-49 | 1.52 (1.36, 1.71) | |
| | 50-59 | 1.62 (1.50, 1.75) | |
| | 60-69 | 1.58 (1.47, 1.71) | |
| | ≥70 | 1.49 (1.33, 1.67) | |
| LDpred2 PRS | <40 | 1.63 (1.35, 1.97) | $3.90Ex10^{-44}$ |
| | 40-49 | 1.44 (1.29, 1.61) | |
| | 50-59 | 1.69 (1.56, 1.84) | |
| | 60-69 | 1.79 (1.66, 1.94) | |
| | ≥70 | 1.39 (1.24, 1.57) | |
| PRS-CSx | <40 | 1.55 (1.30, 1.87) | $1.59x10^{-33}$ |
| | 40-49 | 1.49 (1.34, 1.67) | |
| | 50-59 | 1.73 (1.60, 1.88) | |
| | 60-69 | 1.84 (1.70, 1.99) | |
| | ≥70 | 1.45 (1.29, 1.63) | |
| CT-SLEB | <40 | 1.70 (1.42, 2.05) | $1.60x10^{-3}$ |
| | 40-49 | 1.51 (1.36, 1.69) | |
| | 50-59 | 1.75 (1.52, 1.90) | |
| | 60-69 | 1.93 (1.79, 2.08) | |
| | ≥70 | 1.53 (1.37, 1.72) | |

Abbreviations: OR, odds ratio; CI, confidence interval; CT, clumping and thresholding

*Interaction was evaluated using the likelihood ratio test

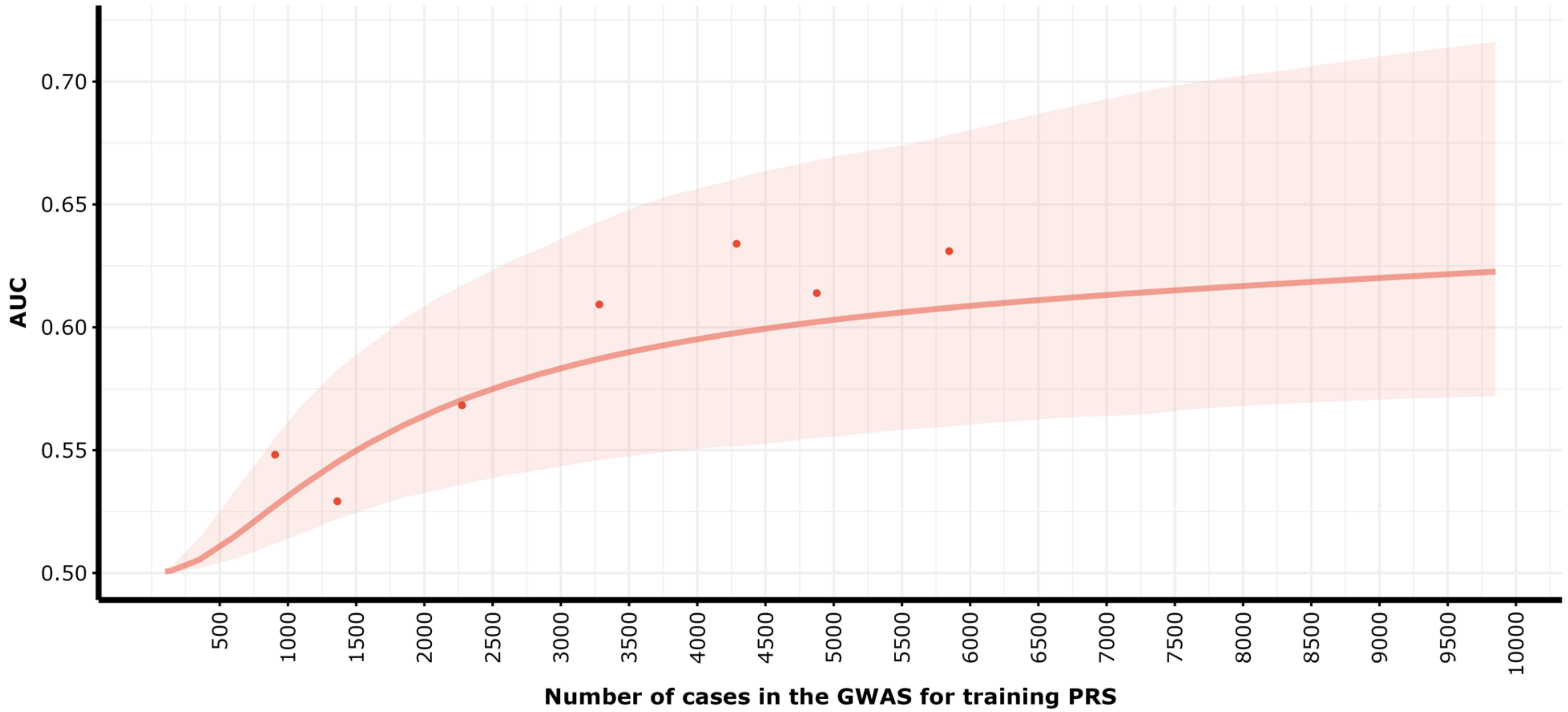


**Figure S1. Observed LDpred2 PRS AUC on seven different case sample sizes and projected LDpred2 PRS AUC of case sample sizes in the range of [500, 10,000] (with case to control ratio of 1:1).** Dots represent observed AUC, the line represents the projected AUC, and the shaded area represents the 95% CI of projected AUC.

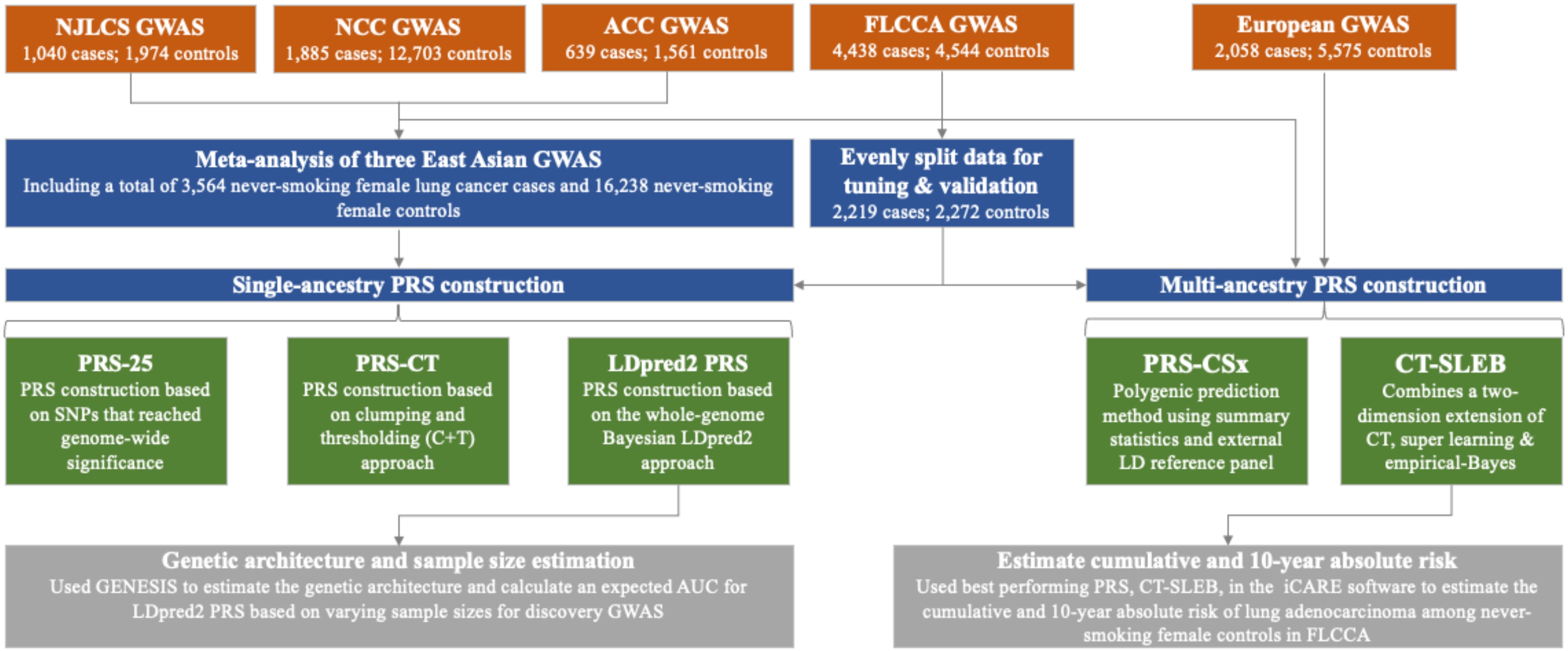


**Figure S2. Overview of data structure, polygenic risk score (PRS) development, validation and application.** Summary statistics from a meta-analysis of East Asian genome-wide association studies (GWAS), including samples from Nanjing Lung Cancer Study (NJLCS), National Cancer Center of Japan (NCC), and the Research Institute and Aichi Cancer Center (ACC) were used to develop single-ancestry PRS using methods such as a simple PRS constructed using 25 SNPs that have previously reached genome-wide significance (i.e., $P<5x10^{-8}$) (PRS-25), a PRS using the clumping and thresholding (CT) methods (PRS-CT), and a PRS using a genome-wide Bayesian-based approach, LDpred2 (LDpred2 PRS). For the multi-ancestry PRS development, we also used summary statistics from European (EUR) GWAS, applying the PRS-CSx method that leveraged genome-wide association summary statistics with a Bayesian continuous shrinkage prior to model SNP effect sizes across populations, as well as CT-SLEB method, which enhances the standard CT methods with a two-dimensional approach to select SNPS for East Asian PRS construction. Tuning and validation of each PRS was conducted in the Female Lung Cancer Consortium in Asian, an independent East Asian individual-level dataset. CT-SLEB PRS was using to estimate 10-year and lifetime cumulative absolute risk, and PRS-CT and PRS-LDpred2 were used for sample size projection.

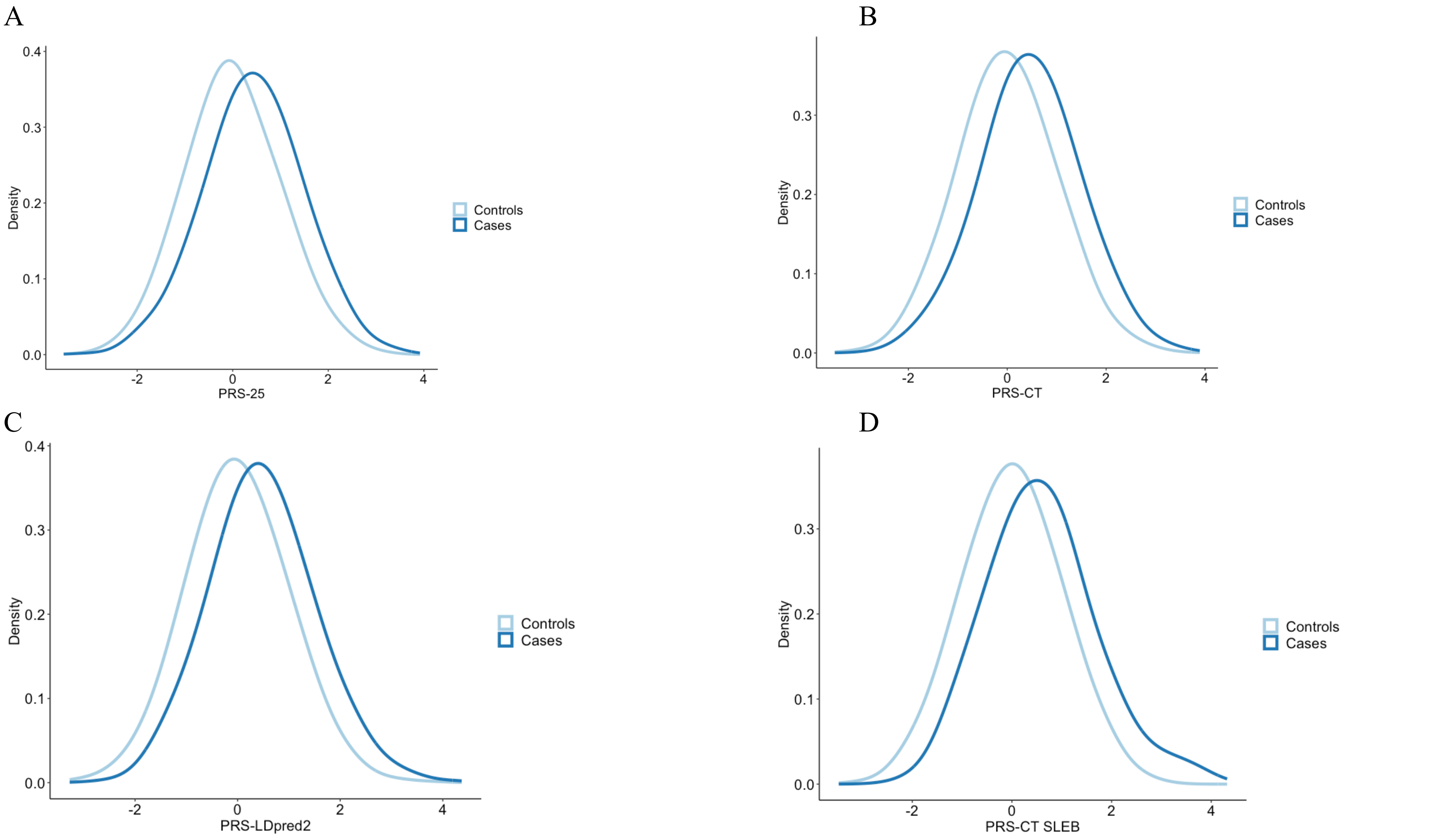


**Figure S3. PRSs distribution in the Female Lung Cancer Consortium in Asia**

| | PRS-25 | | | PRS-CT | | | LDpred2 PRS | | | CT-SLEB | | |
|---|---|---|---|---|---|---|---|---|---|---|---|---|
| | Cases | Controls | Total | Cases | Controls | Total | Cases | Controls | Total | Cases | Controls | Total |
| **Mean** | 0.43 | 0 | 0.21 | 0.34 | 0 | 0.17 | 0.51 | 0 | 0.25 | 0.58 | 0 | 0.29 |
| **SD** | 1.03 | 1 | 1.04 | 1.03 | 1 | 1.03 | 1.08 | 1 | 1.07 | 1.11 | 1 | 1.09 |

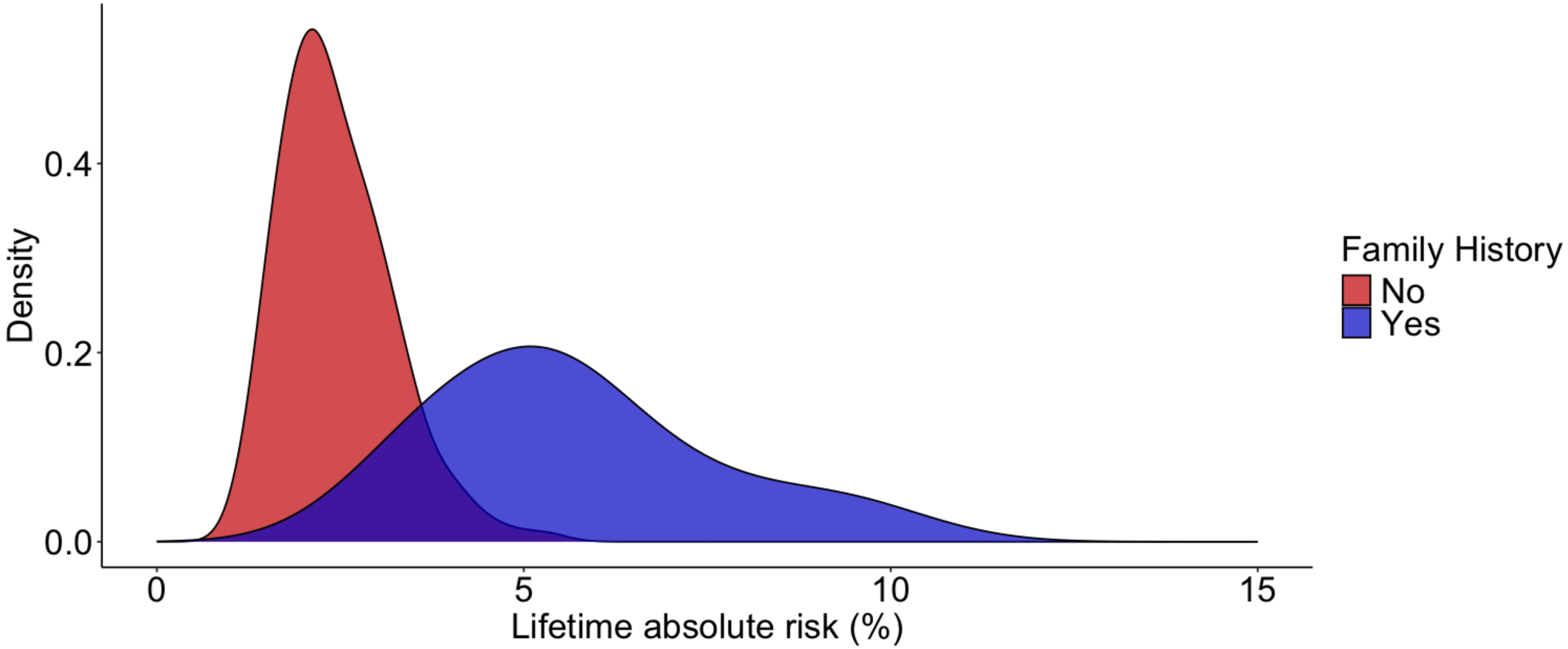


**Figure S4. Lifetime cumulative absolute risk of developing lung adenocarcinoma by family history**. Lifetime (age 30-80) absolute risk of developing lung adenocarcinoma in never-smoking East Asian women stratified by first-degree family history of lung cancer. Absolute risks were calculated using iCARE package[42], based on Taiwan's age specific incidence and mortality data, and the relative risks of the PRS and family history, as described in the Methods section.

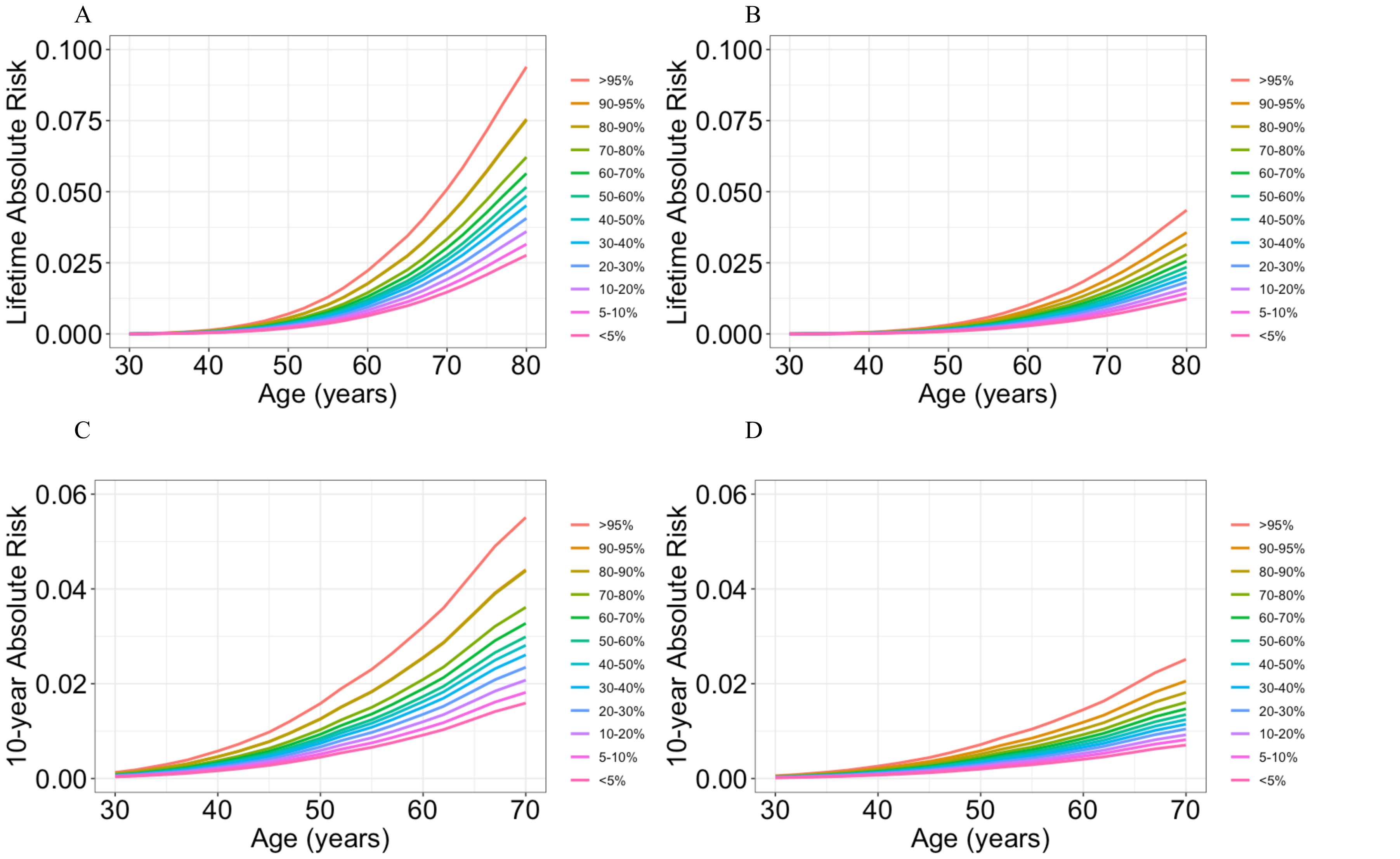


**Figure S5. Lifetime cumulative and 10-year absolute risk of developing lung adenocarcinoma by family history in the Female Lung Cancer Consortium in Asia.** Lifetime (age 30-80) cumulative risk with (A) and without (B) family history, and 10-year absolute risk with (C) and without (D) family history of developing lung adenocarcinoma in never-smoking women in East Asia by percentiles of the CT-SLEB polygenic risk score (PRS). Absolute risks were calculated using the iCARE package, based on Taiwan's age-specific incidence and mortality data, and the PRS relative risks, as described in the Methods section.
